# Identifying profiles, trajectories, burden, social and biological factors in 3.3 million individuals with multimorbidity in England

**DOI:** 10.1101/2025.05.24.25326850

**Authors:** Yu Liu, Clare Bankhead, Cynthia Wright Drakesmith, Catherine Pope, David Gonzalez-Chica, Subhashisa Swain, CoMPuTE, Carl Heneghan, Rafael Perera-Salazar, Tingting Zhu

**Author notes:** A list of authors and their affiliations appears at the end of the paper.

## Abstract

Multimorbidity, the co-occurrence of multiple chronic conditions in an individual, has become a global health challenge affecting populations in high-income and low- to middle-income countries. Despite its increasing prevalence, critical gaps remain in understanding its progression, burden, and determinants to better guide prevention and treatment. Here, by leveraging linked primary care, hospitalisation, and mortality records from 3.3 million individuals with multimorbidity in England, we conducted a longitudinal cohort study to characterise multimorbidity across multiple dimensions, including condition profiling, progression trajectories, healthcare burden, and associated social and biological factors. Specifically, we identified 21 distinct multimorbidity profiles in males and 18 in females, uncovering life-course progression pathways. We assessed the differential burden of these profiles on mortality and hospitalisation. The study also highlights how social inequalities shape distinct patterns of multimorbidity. Furthermore, by developing an interpretable machine learning framework, we identified key biological markers associated with specific multimorbidity profiles. Together, these results offer valuable insights to inform prevention strategies, public health initiatives and potential interventions aimed at mitigating the growing burden of multimorbidity.

## Introduction

Multimorbidity, the co-occurrence of two or more chronic conditions in an individual, threatens individual health and overwhelms healthcare systems^1,2^. Globally, more than one-third of adults are affected^3,4^, and in England alone, annual healthcare costs per patient increase from £3,000 for those with a single condition to £8,000 for those with three or more conditions^5^. Despite the growing burden, most clinical guidelines remain focused on single conditions, making them inadequate for people with multiple mental and physical health conditions, who are generally associated with poorer quality of life, shorter life expectancy, greater functional difficulties, and increased use of healthcare^1^. In addition, the complexity of multimorbidity extends beyond the simple accumulation of conditions, as its mechanisms vary quantitatively and qualitatively across sexes, stages of life, and a spectrum of social and biological factors^1,6–10^, highlighting the urgent need to better understand multimorbidity to enable more effective prevention, earlier detection, and improved clinical management.

As the accumulation of conditions is a well-recognised characteristic of multimorbidity, several studies have measured it by counting the number of conditions per individual^11–13^. However, these studies lack information on the composition and interplay of conditions within multimorbidity. Motivated by the observation that individuals with common conditions tend to cluster in clinical practice and epidemiological studies^14^, various methods have been used to identify multimorbidity profiles in cross-sectional studies^15,16^, with latent class analysis (LCA)^17^ emerging as the most widely used approach^7,18^. Several clinically meaningful profiles have been identified, such as cardiovascular and cardiometabolic profiles^19,20^. Although these studies have improved our understanding of generic patterns in multimorbidity, their crosssectional design fails to capture the longitudinal progression.

Taking advantage of the digitalisation of healthcare systems and the accumulation of substantial longitudinal data from electronic health records (EHRs), some studies have investigated multimorbidity trajectories^21–24^. Evidence indicates substantial transitions from profiles characterised by cardiovascular risk factors such as diabetes and hypertension, to those dominated by explicit cardiovascular conditions such as heart attack and stroke, reflecting the dynamic nature of multimorbidity^25^. However, most studies followed individuals for less than 15 years^26^. Such time frames capture only partial life stages, such as early adulthood (e.g., 25–45 years), middle adulthood (e.g., 45–65 years) or late adulthood (e.g., 65–85 years)^27^. Therefore, longitudinal studies that span the full life course are essential to fully characterise multimorbidity trajectories, and understand their cumulative burden on individual quality of life over time.

In parallel, the role of social and biological factors in shaping multimorbidity has been increasingly recognised^1,6–8^. Socioeconomic inequalities, for example, have been shown to significantly influence multimorbidity, with disadvantaged groups experiencing earlier onset and faster accumulation of chronic conditions^28–30^. Despite these insights, the specific contributions of social factors to the emergence and persistence of different multimorbidity profiles remain poorly understood^20^. From a biological perspective, while clinical markers such as high body mass index (BMI) have been linked to multimorbidity development^31^, few studies have systematically examined multiple routinely collected clinical measures^10,32^, hindering a deeper understanding of the underlying biological factors.

Taken together, despite growing recognition of multimorbidity’s complexity, no study has systematically characterised its composition, progression, and impact, alongside underlying social and biological factors, within a large population-based cohort to inform more effective preventive strategies and targeted interventions^8,33^. Motivated by this gap, we examined key aspects of multimorbidity throughout the life course of 3.3 million individuals in England using the Clinical Practice Research Datalink (CPRD) data^34^, a representative sample of the English primary care population^35,36^. The study was structured around five objectives: (1) We identified multimorbidity profiles using LCA and hierarchical clustering, stratified by age and sex; (2) We characterised life-course multimorbidity trajectories throughout the study population; (3) We assessed the burden of multimorbidity in terms of both mortality and hospitalisation; (4) We analysed the association between multimorbidity and social factors, including socioeconomic deprivation, ethnicity and geography; (5) We investigated the biological factors underlying multimorbidity, developing an interpretable machine learning framework to identify clinically relevant markers for specific multimorbidity profiles.

## Results

### Data overview and study design

Using routinely collected EHR data from CPRD^34^, we derived the longitudinal diagnoses of 18 chronic conditions in 6,671,245 individuals. These conditions represent the commonly reported chronic conditions in the general population^37–40^, across mental, respiratory, metabolic, cardiovascular, neurological, and other systems (Methods). Individuals entered the study at birth and were followed until death or study exit, with follow-up period ended no later than 31 December 2019. Among the study cohort, 3,314,652 individuals developed multimorbidity (i.e. two or more of the investigated chronic conditions) at the time of death or exit, with an overall prevalence of 49.69% (95% confidence interval [CI]: 49.65–49.72%). These individuals made up the primary study cohort, with a median duration of 68 years (interquartile range [IQR]: 50–81 years) from birth to death or exit, and a median year of birth of 1950 (IQR: 1936– 1968; Supplementary Fig. S1). Table 1 summarises the basic characteristics of this group. For comparison, we identified 1,226,245 individuals from the study cohort who remained free of all 18 conditions throughout the study period, referred to as the healthy cohort. The remaining 2,130,348 individuals developed one of the 18 conditions during the study period. For clarity, we refer to some of selected conditions using abbreviations: coronary heart disease (CHD), chronic obstructive pulmonary disease (COPD), and serious mental illnesses (SMI).

**Table 1:**
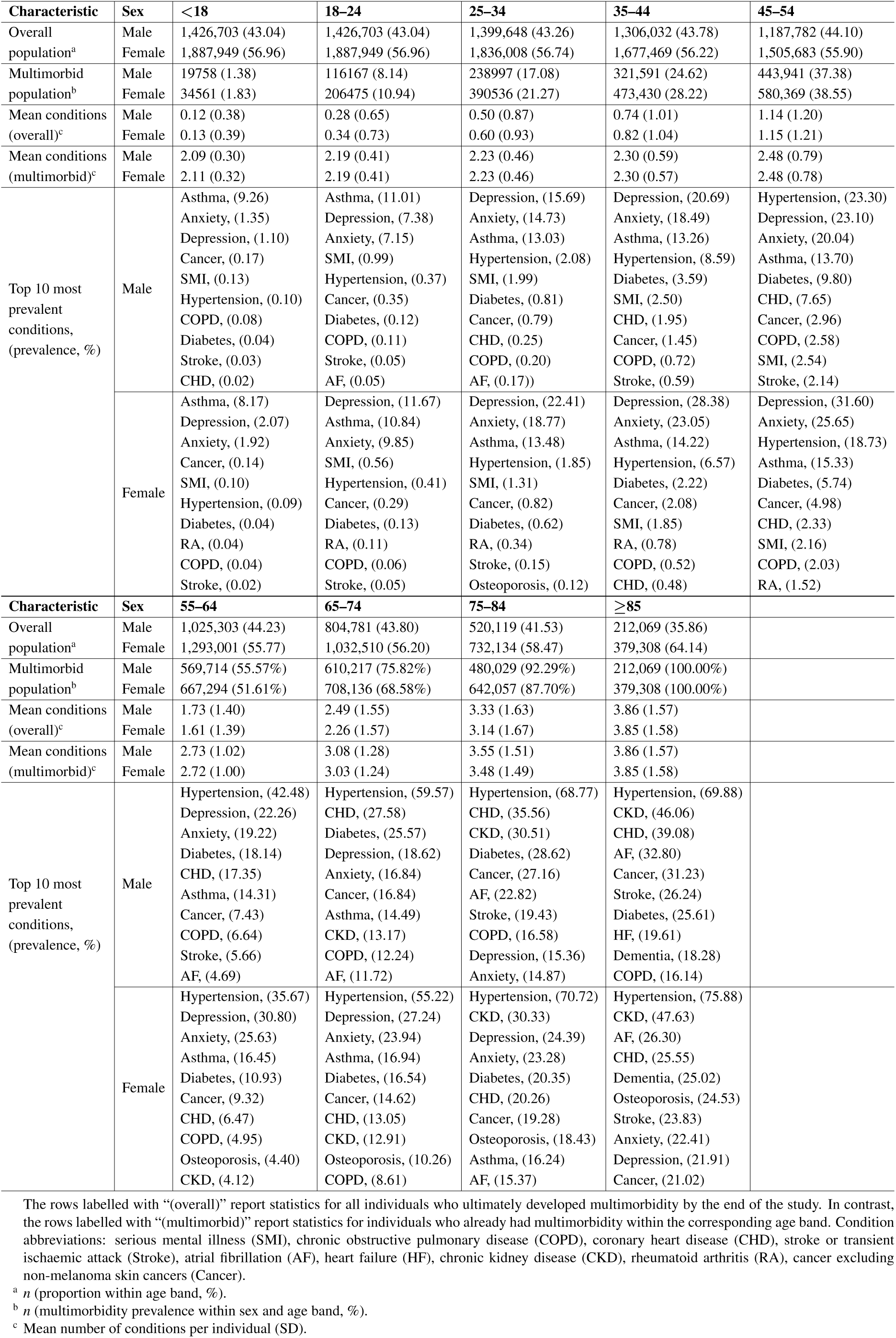
Characteristics of each stratum in the primary study cohort.

The overall framework of our analysis is described in Fig. 1. To capture multimorbidity progression over time, we divided each individual’s clinical history into nine age bands (*<*18, 18–24, 25–34, 35–44, 45–54, 55–64, 65–74, 75–84, ≥85) and stratified analyses by sex. Within each age band, LCA^17^ was applied to identify clusters of individuals with similar diagnostic profiles (Fig. 1a). To identify generic multimorbidity profiles across the life course, clusters from different age bands were merged using hierarchical clustering based on condition prevalence and exclusivity patterns (Fig. 1b and Supplementary Fig. S2–S4). Life-course multimorbidity trajectories were reconstructed by linking individuals’ profiles across successive age bands (Fig. 1c). Using 847,048 death records and 33,950,515 hospitalisation records from the Office for National Statistics (ONS)^41^ and Hospital Episode Statistics (HES)^42^, we assessed the burden of multimorbidity in terms of mortality and hospitalisation across profiles and age bands (Fig. 1d). We then analysed the association between multimorbidity profiles and various social factors (Fig. 1e and Supplementary Table S1). For socioeconomic deprivation, individuals were linked to quintiles of the 2019 English Index of Multiple Deprivation (IMD) as a measure of relative deprivation^28^. For ethnicity, individuals were grouped by White, Black, South Asian, Mixed, and Other^12^. For geography, each individual was assigned to one of nine regions in England. Using 1,064,737,538 records covering 45 clinical markers from CPRD, we developed an interpretable machine learning framework that combines XGBoost^43^ with a novel reference-adjusted Shapley additive explanation (SHAP) method^44,45^ to identify clinically meaningful markers associated with each multimorbidity profile (Fig.1f and Methods). The complete marker list appears in Supplementary Table S2.

**Figure 1:**
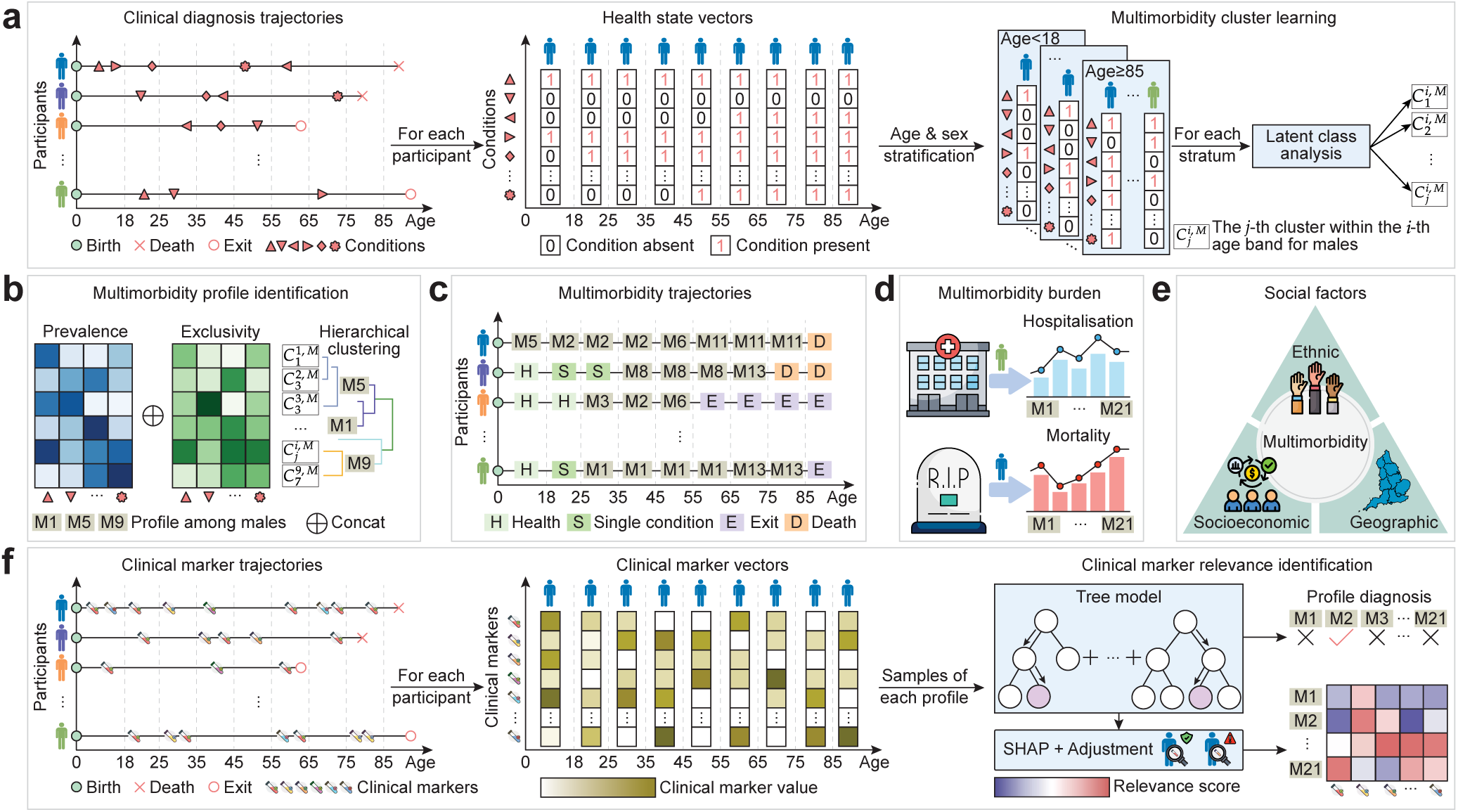
Study overview. **a,** Clinical diagnosis trajectories were extracted for participants over the life course, stratified by sex and age band. The example shown is for males. Trajectories were encoded as health state vectors representing the presence of specific conditions at each age band. Latent class analysis was applied within each stratum to identify multimorbidity clusters (e.g., *C_j_^i,M^* denotes the *j*-th cluster within the *i*-th age band for males). **b,** To capture consistent multimorbidity patterns across age bands, hierarchical clustering was performed based on condition prevalence and exclusivity within the cluster. Condition prevalence was defined as the proportion of individuals within a cluster who had a given condition, whereas condition exclusivity was defined as the proportion of individuals with a specific condition in a given age band who belonged to that particular cluster. This yielded generic multimorbidity profiles (e.g., M1, M5, M9). **c,** Individual multimorbidity trajectories were reconstructed by mapping transitions across profiles over the life course. **d,** The burden associated with each profile was quantified using mortality, hospitalisation rate (mean annual hospitalisations per individual), and hospitalisation prevalence (proportion of individuals experiencing at least one hospitalisation). **e,** Descriptive analyses examined associations between multimorbidity and social factors, including socioeconomic deprivation, ethnicity, and geographic regions. **f,** An interpretable machine learning framework was developed to examine the association between multimorbidity and biological factors. Clinical marker trajectories over the life course were extracted and encoded into clinical marker vectors, stratified by sex and age band. A tree-based classifier XGBoost was trained to predict individual multimorbidity profiles from these vectors. A Shapley value–based explainer with novel reference-based adjustment was applied to identify clinically relevant markers for each profile.

### Multimorbidity profiles

Throughout the life course, 21 multimorbidity profiles for males and 18 for females were identified in the primary study cohort (Fig. 2). Detailed numerical results are provided in Supplementary Fig. S5 and Table S3–S4, and the naming convention for profiles is provided in Supplementary Note S1.2.

**Figure 2:**
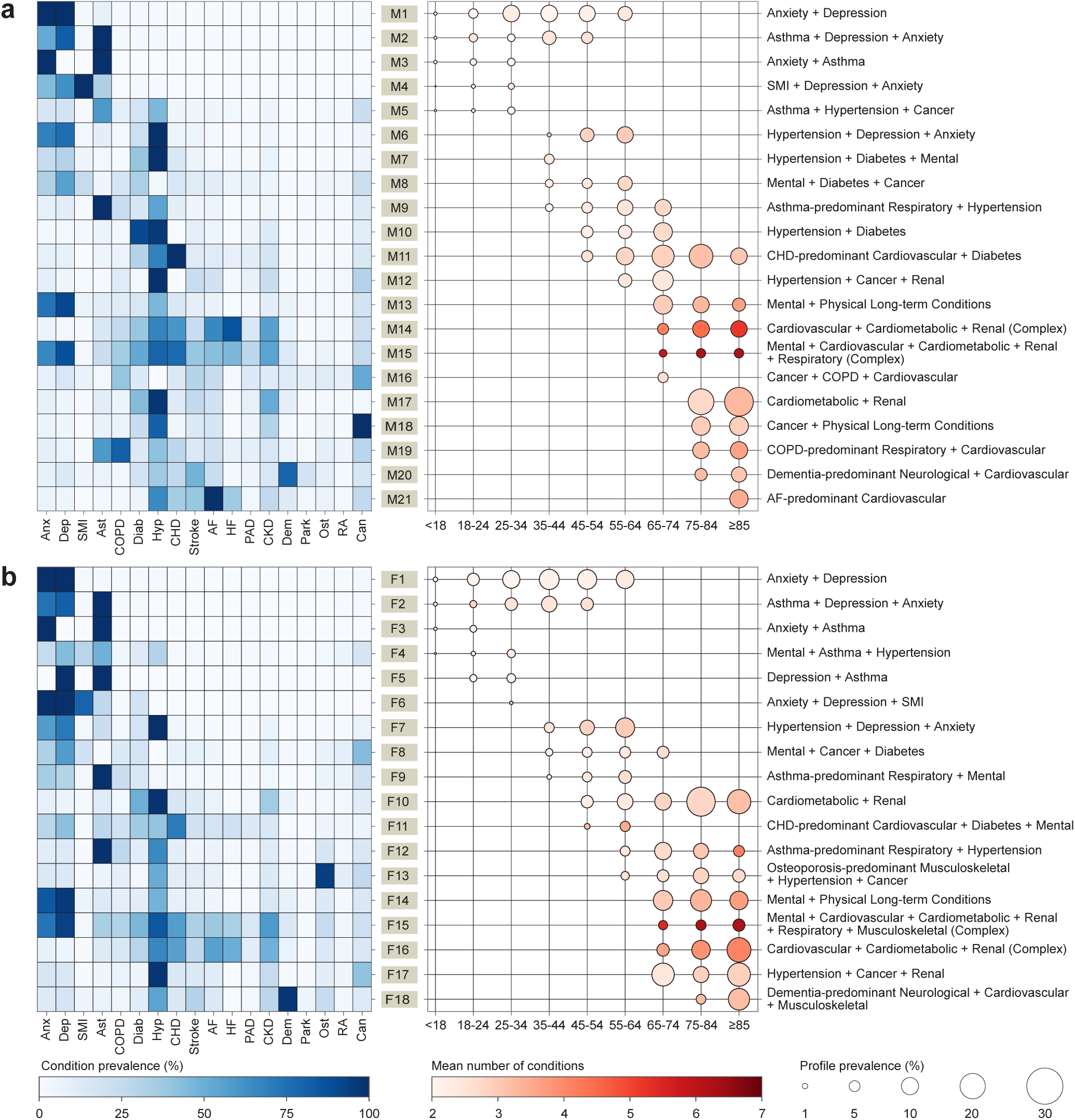
Characteristics of the identified multimorbidity profiles for a, Males and b, Females. In each panel, the left part presents a heatmap of condition prevalence within each profile, with darker blue shades indicating higher prevalence (numerical results are provided in Supplementary Fig. S5). The middle part presents a bubble plot, where the size of circles corresponds to the prevalence for a profile across an age band (numerical results are provided in Supplementary Table S3–S4). The colour intensity reflects the mean number of conditions per individual, with darker red shades indicating a higher number of conditions. Profile labels are positioned between the two plots, and the right part lists the name of each profile, determined using the convention outlined in Supplementary Note S1.2. Conditions listed in the profile names are ordered by prevalence, and those with a mean number of conditions exceeding four are annotated as “Complex”. Condition abbreviations: anxiety (Anx), depression (Dep), serious mental illness (SMI), asthma (Ast), chronic obstructive pulmonary disease (COPD), diabetes (Diab), hypertension (Hyp), coronary heart disease (CHD), stroke or transient ischaemic attack (Stroke), atrial fibrillation (AF), heart failure (HF), peripheral arterial disease (PAD), chronic kidney disease (CKD), osteoporosis (Ost), rheumatoid arthritis (RA), cancer excluding non-melanoma skin cancers (Can).

The multimorbidity profiles showed clear age-related trends that became increasingly more prevalent and complex with ageing, in terms of both composition and accumulation of conditions. In early life (*<*35 years), profiles were predominantly characterised by mental health conditions, often co-occurring with asthma (M1–M5 in males and F1–F6 in females). In midlife (35–64 years), physical conditions such as hypertension, diabetes, and cancer became more prominent (M6–M12, F7–F13). In later life (≥65 years), newly emerging profiles reflected increased complexity and multisystem involvement, especially in the cardiovascular, cardiometabolic, renal, and mental health domains (M13–M21, F14–F18).

Complex multimorbidity profiles, defined as those with more than four conditions on average^6^, were identified in older adults. In the ≥85 age band, males in M14 (Cardiovascular + Cardiometabolic + Renal, Complex) and M15 (Mental + Cardiovascular + Cardiometabolic + Renal + Respiratory, Complex) had average condition counts of 5.86 (SD: 1.34) and 7.03 (SD: 1.30), respectively, while females in F16 (Cardiovascular + Cardiometabolic + Renal, Complex) and F15 (Mental + Cardiovascular + Cardiometabolic + Renal + Respiratory + Musculoskeletal, Complex) had averages of 4.53 (SD: 1.48) and 7.02 (SD: 1.33). Mental health conditions were over-represented in M15 and F15, with depression affecting more than 85% of the individuals in both profiles, compared to less than 20% in M14 and F16. In contrast, M14 and F16, while also complex, showed a higher prevalence of heart failure and atrial fibrillation. The prevalence of heart failure was 87.11% (95% CI: 86.90–87.31%) in M14 and 50.22% (95% CI: 50.01–50.43%) in F16, compared to 46.04% (95% CI: 45.54–46.54%) in M15 and 33.42% (95% CI: 33.08–33.76%) in F15.

Several multimorbidity profiles exhibited long persistence—appearing repeatedly across successive age bands. The mental health-only profiles (M1 and F1: Anxiety + Depression) were observed across six age bands under age 65, highlighting their widespread impact across a broad population. Among profiles dominated by physical conditions, F10 (Cardiometabolic + Renal) in females and M11 (CHD-predominant Cardiovascular + Diabetes) in males were the most persistent, each spanning five consecutive age bands from age 45 and above.

Sex differences in multimorbidity patterns were evident, with males being more frequently included in cardiovascular, respiratory, and cancer-related profiles. Six cardiovascular-related profiles were identified in males (M11, M14, M15, M19–M21) compared to four in females (F11, F14, F15, F18). Although both sexes had a CHD-predominant profile (M11, F11), M11 was more prevalent and persisted throughout a broader age range, while F11 was observed only in two age bands (45–64 years). Although asthma-predominant respiratory profiles were common in both sexes (M9, F9, F12), severe respiratory conditions such as COPD were predominantly observed in males: M19 (COPD-predominant Respiratory + Cardiovascular) showed a COPD prevalence of 81.37% (95% CI: 81.09–81.70%). Similarly, cancer appeared more prominently in male profiles, with a prevalence of 51.70% (95% CI: 51.13– 52.27%) in M16 (Cancer + COPD + Cardiovascular) and 100% in M18 (Cancer + Physical Long-term Conditions). Unlike males, females had a higher prevalence of mental, neurological, and musculoskeletal profiles. For example, F14 (Mental + Physical Long-term Conditions) had a prevalence of 15.54% (95% CI: 15.46–15.62%) in ages 75–84, compared to 9.91% (95% CI: 9.83–9.99%) of the comparable male profile M13 (Mental + Physical Longterm Conditions). Dementia-predominant neurological profiles were also more prevalent in females (F18: 15.76%, 95% CI: 15.65–15.88% in ≥85 years) than males (M20: 8.07%, 95% CI: 7.95–8.18%), and often involved musculoskeletal conditions. A female-specific profile, F13 (Osteoporosis-predominant Musculoskeletal + Hypertension + Cancer), spanned four age bands from age 55 and had an osteoporosis prevalence of 94.87% (95% CI: 94.76–94.97%).

### Multimorbidity trajectories

Longitudinal transitions between multimorbidity profiles across age bands were observed (Fig. 3 and Supplementary Fig. S6). The Sankey diagrams illustrate that the population with multimorbidity increased with age, peaking in the 65–74 age band as individuals accumulated conditions. This was followed by a decline in the older age bands due to mortality and exit from the study. Several distinct progression pathways were identified.

**Figure 3:**
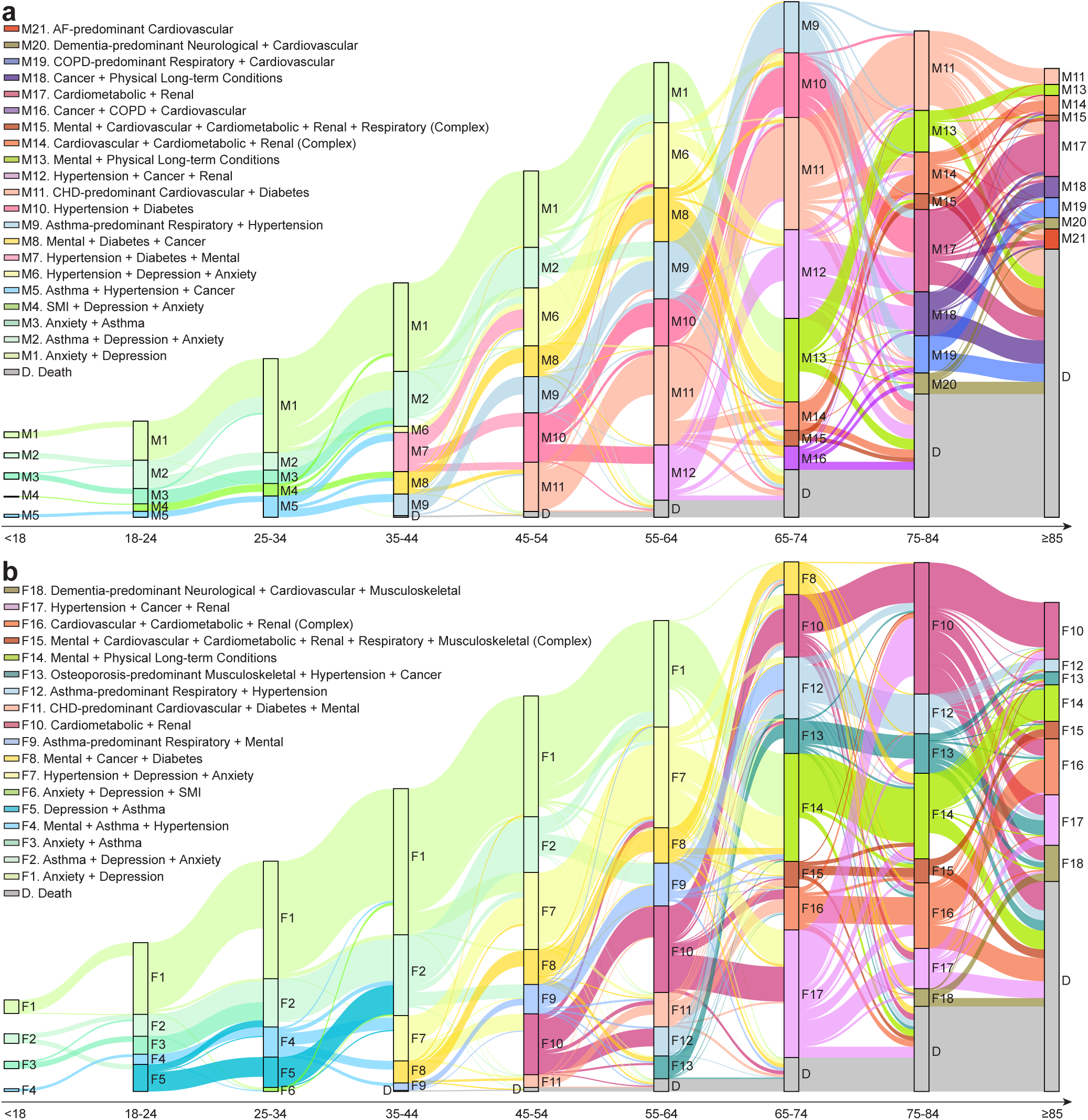
Multimorbidity trajectories over the life course for a, Males and b, Females. The Sankey diagrams illustrate transitions of the 3.3 million individuals in the primary study cohort between multimorbidity profiles across different age bands. For each panel, the height of each bin is proportional to the number of individuals within the corresponding profile. Each coloured flow represents the transition of individuals from one profile to another, where the thickness of the flow is proportional to the number of individuals in the transition. Transitions from the same source profile are shown in the same colour. The common profiles across sexes are represented using the same colour, and profiles with similar condition compositions use similar hues. For clarity, transitions from individuals with no or single conditions into multimorbidity profiles are omitted, and the complete trajectories for the whole population are presented in Supplementary Fig. S6. An interactive demo of this figure is available for males and females.

In both sexes, mental health–predominant profiles often represented early multimorbidity, and individuals within these profiles progressed to more complex profiles incorporating age-related physical conditions. The mental health-only profiles (M1 and F1: Anxiety + Depression) frequently transitioned into profiles that added hypertension and diabetes. For example, between ages 45–64, a subset of individuals from M1/F1 moved to M6/F7 (both Hypertension + Depression + Anxiety). In the 65–74 age band, M13 and F14 (both Mental + Physical Longterm Conditions) emerged, with 41.29% of males and 57.93% of females originating from M1 and F1, an additional 35.62% of males and 29.37% of females from M6 and F7, and the remaining individuals contributed from M8 (Mental + Diabetes + Cancer) and F8 (Mental + Cancer + Diabetes), respectively.

For cardiovascular-predominant trajectories, direct transitions from healthy or single-condition states into M11 (CHD-predominant Cardiovascular + Diabetes) were common. 72.32% and 64.68% of individuals entered M11 from these groups in the 45–54 and 55–64 age bands, respectively. Subsequently, M11 served as a key precursor to more complex profiles such as M14 (Cardiovascular + Cardiometabolic + Renal, Complex), contributing 42.25% and 31.22% of its population in the 65–74 and 75–84 age bands, respectively. A similar pattern was observed in females, with transitions from F11 (CHD-predominant Cardiovascular + Diabetes + Mental) to F16 (Cardiovascular + Cardiometabolic + Renal, Complex).

For cardiometabolic- and renal-predominant trajectories, notable sex-specific patterns were observed. In males, a considerable proportion of individuals transitioned from healthy or single-condition states to M10 (Hypertension + Diabetes), which persisted across three age bands (45–74 years), and M12 (Hypertension + Cancer + Renal), present at ages 55–74. These two profiles served as key intermediates in the development of M17 (Cardiometabolic + Renal) in the 75–84 age band, together accounting for 70.95% of its population. They also preceded M18 (Cancer + Physical Long-term Conditions), contributing 79.70% of its population. In females, a similar pathway was observed involving F10 (Cardiometabolic + Renal) and F17 (Hypertension + Cancer + Renal), both of which included a substantial number of individuals transitioning from healthy or single-condition states, and 45.50% of individuals transitioned from F17 to F10 in the 75–84 age band. Both contributed to the formation of F16 (Cardiovascular + Cardiometabolic + Renal, Complex).

Distinct sex differences were evident in respiratory-predominant trajectories. In males, M9 (Asthma-predominant Respiratory + Hypertension), observed across four age bands (35–74 years), often served as a precursor to M19 (COPD-predominant Respiratory + Cardiovascular) in the 75–84 age band, with 43.53% of individuals making this transition. In females, respiratory progression was more closely intertwined with mental health conditions. F5 (Depression + Asthma), present at ages 18–34, transitioned almost entirely (96.14%) into F2 (Asthma + Depression + Anxiety) in the 35–44 age band due to the accumulation of mental health conditions. Between ages 45–64, F2 contributed over 45% of individuals to F9 (Asthma-predominant Respiratory + Mental). Subsequently, 41.71% of the individuals in F9 transitioned to F12 (Asthma-predominant Respiratory + Hypertension), in which hypertension was highly prevalent (65.31%, 95% CI: 65.12–65.51%).

In addition, we examined the number of unique multimorbidity profiles each individual experienced (Supplementary Fig. S7). Among the 3.3 million individuals in the primary study cohort, 58.53% of males and 58.43% of females stayed in a single profile across all age bands, while 35.40% of males and 34.74% of females experienced two distinct profiles. The remaining 6.07% of males and 6.83% of females transitioned through three or more profiles. On average, males experienced 1.48 (SD: 0.62) profiles, and females experienced 1.49 (SD: 0.64) profiles during the study period.

### Burden of multimorbidity

The burden associated with multimorbidity was assessed using three indicators: mortality (Fig. 4a,b), hospitalisation rate (mean annual hospitalisations per individual; Fig. 4c,d), and hospitalisation prevalence (proportion of individuals experiencing at least one hospitalisation; Supplementary Fig. S8), stratified by multimorbidity profile, sex, and age. Results from the healthy cohort were included as a reference. Across all indicators, the burden of multimorbidity increased progressively with age. Profiles dominated by physical conditions were generally associated with greater burden than those primarily driven by mental health conditions, with complex multimorbidity profiles imposing the highest burden overall.

**Figure 4:**
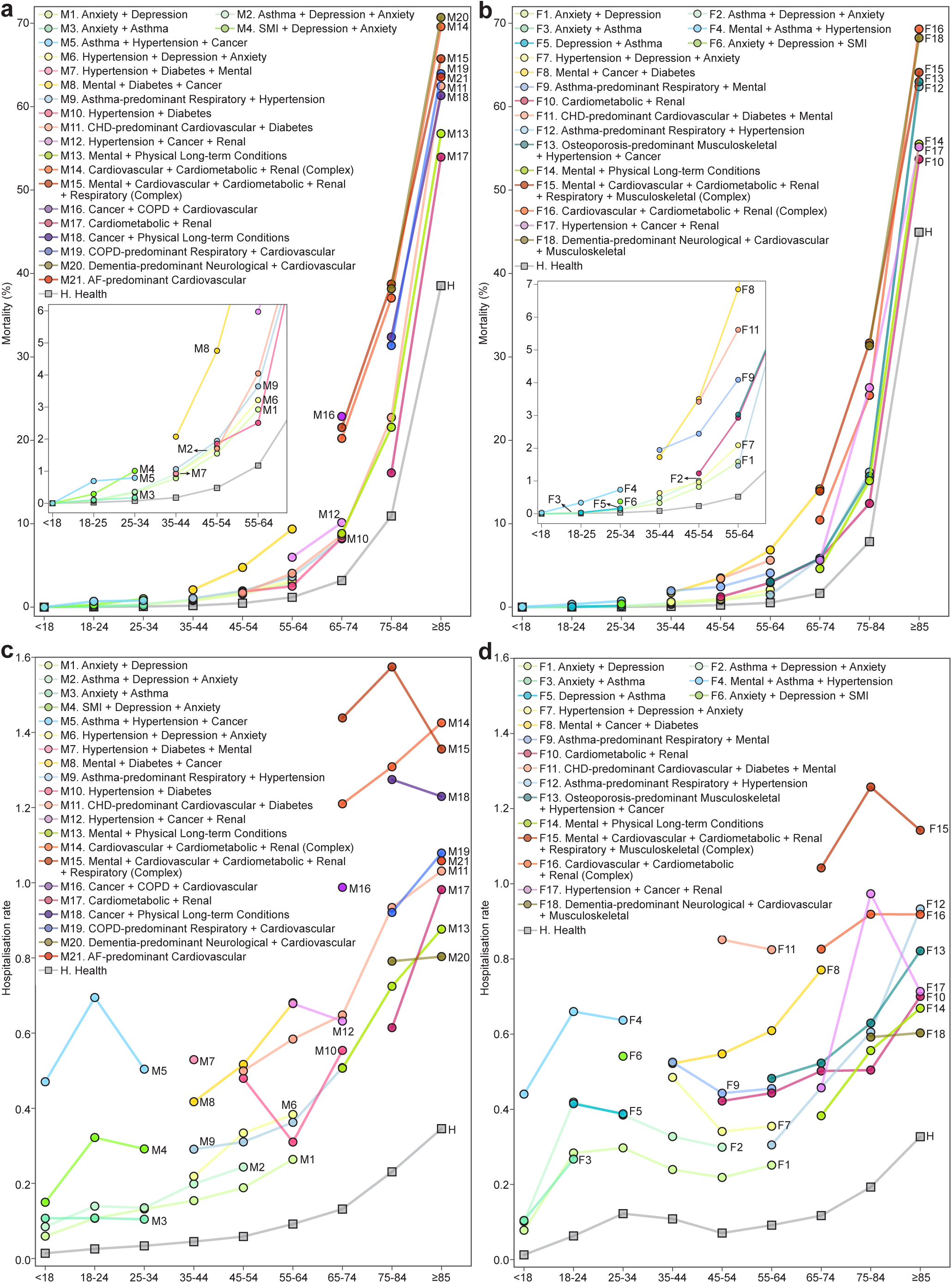
Multimorbidity burden over the life course. **a, b,** Mortality across multimorbidity profiles and age bands in males and females, respectively. Insets show a magnified view of the age bands below 65 years. **c, d,** Hospitalisation rate across multimorbidity profiles and age bands in males and females, respectively. For each panel, each line represents a multimorbidity profile, with values calculated within profile-specific cohorts for each age band. Mortality was defined as the proportion of individuals who died within each profile and age band. Hospitalisation rate was defined as the average number of hospitalisations per person-year within each profile for an age band. The healthy cohort is shown in grey for comparison. The colour coding follows the same scheme as in Fig. 3. More results on hospitalisation prevalence are provided in Supplementary Fig. S8.

Mortality remained low across profiles before age 65, as compared to the levels observed in the healthy cohort. Notable exceptions were M8 (Mental + Diabetes + Cancer) and F8 (Mental + Cancer + Diabetes), with mortality respectively reaching 9.36% (95% CI: 9.15–9.57%) and 6.85% (95% CI: 6.64–7.06%) in the 55–64 age band. After age 65, mortality rose sharply, and stratified into three tiers in the ≥85 age band. The highest mortality (∼70%) was observed in neurological-predominant profiles (M20: Dementia-predominant Neurological + Cardiovascular, F18: Dementia-predominant Neurological + Cardiovascular + Musculoskeletal) and complex profiles (M14 and F16: Cardiovascular + Cardiometabolic + Renal, Complex). In contrast, mortality was lower (∼55%) in mental health profiles among older individuals (M13 and F14: Mental + Physical Long-term Conditions) and cardiometabolic profiles (M17 and F10: Cardiometabolic + Renal), with remaining profiles falling between 60–65%. Mortality in the healthy cohort in the same age band was 38.54% (95% CI: 37.50–39.60%) for males and 44.95% (95% CI: 44.06–45.84%) for females. Although mortality across multimorbidity profiles was similar between the sexes, this gap relative to the healthy cohort suggests a disproportionate mortality burden on older males.

Interestingly, although M15 (Mental + Cardiovascular + Cardiometabolic + Renal + Respiratory, Complex) and F15 (Mental + Cardiovascular + Cardiometabolic + Renal + Respiratory + Musculoskeletal, Complex) had the highest average number of conditions (7.03 and 7.02, respectively), their mortality was lower than M14 and F16, which had fewer conditions on average (5.86 and 4.53, respectively) but were heavily burdened by cardiovascular conditions such as heart failure and atrial fibrillation. For example, in the ≥85 age band, mortality was 65.75% (95% CI: 64.60–66.88%) in M15, versus 69.60% (95% CI: 68.96–70.23%) in M14.

Hospitalisation rates followed a similar age-related trend as mortality, but varied by sex and profile. In the 18–24 age band, profiles involving hypertension, cancer, and SMI (M4, M5, F5, F6) were associated with elevated hospitalisation rates. Females exhibited higher hospitalisation rates between ages 25–44 due to pregnancy-related care, whereas males showed consistently higher rates in older age bands. Between ages 45–64, cardiovascular-predominant profiles imposed a higher burden on females. For example, F11 (CHD-predominant Cardiovascular + Diabetes + Mental) had a hospitalisation rate of 0.85 (95% CI: 0.78–0.92) in the 45–54 age band, substantially higher than its male counterpart M11 (0.50, 95% CI: 0.47–0.53). In later life, unlike mortality trends, the complex profiles M15 and F15 exhibited the highest hospitalisation rates. M15 and F15 peaked at 1.57 (95% CI: 1.48–1.67) and 1.25 (95% CI: 1.21–1.30), respectively, in the 75–84 age band, while M14 and F16 showed lower rates of 1.31 (95% CI: 1.26–1.35) and 0.92 (95% CI: 0.89–0.94), respectively. Notably, neurological-predominant profiles (M20, F18) exhibited relatively low hospitalisation rates despite high mortality. Similar trends were observed for hospitalisation prevalence (Supplementary Fig. S8), supporting the consistency of the results across different healthcare burden indicators.

### Social factors with multimorbidity

We also examined the multimorbidity profiles in relation to socioeconomic deprivation, ethnicity, and geographic region in England (Fig. 5).

**Figure 5:**
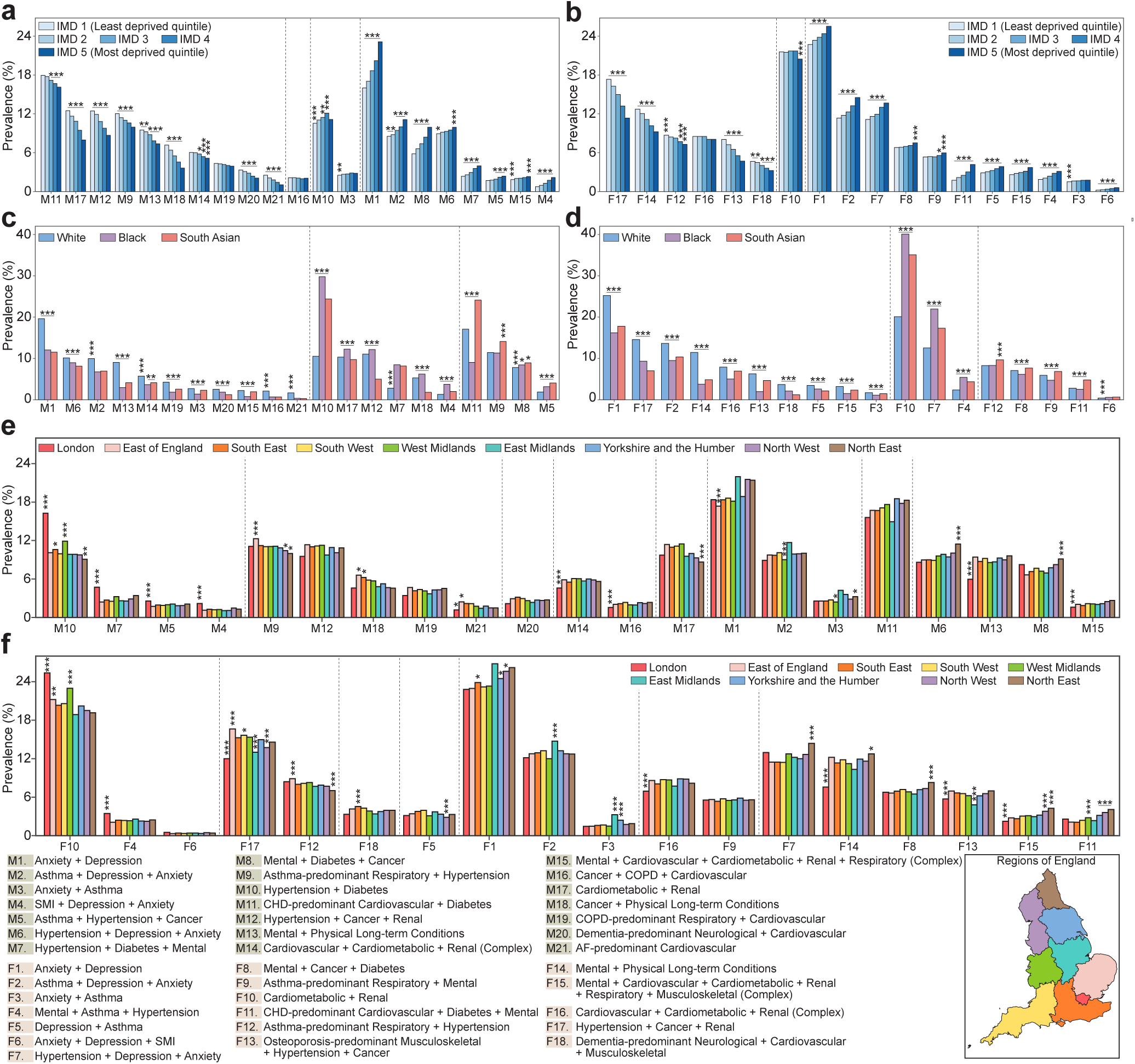
Association between multimorbidity and social factors. **a, b,** Prevalence of multimorbidity profiles across IMD quintiles, from least deprived (IMD 1) to most deprived (IMD 5), for males and females, respectively. For each profile, the prevalence across IMD quintiles is shown. Profiles (i.e., sets of five bars) are sorted according to the IMD quintile with the highest prevalence within each profile. If multiple profiles share the same leading IMD quintile, they are further ordered by descending prevalence within that quintile. Vertical dashed lines separate profiles by the leading IMD quintile. **c, d,** Prevalence of multimorbidity profiles across ethnic groups (White, Black, and South Asian), for males and females, respectively. For each profile, the prevalence across ethnic groups is shown. Profiles (i.e., sets of three bars) are sorted according to the ethnic group with the highest prevalence within each profile. If multiple profiles share the same leading ethnic group, they are further ordered by descending prevalence within that group. Vertical dashed lines separate profiles by the leading ethnic group. **e, f,** Prevalence of multimorbidity profiles across nine regions of England, for males and females, respectively. For each profile, the prevalence across regions is shown. Profiles (i.e., sets of nine bars) are sorted according to the region with the highest prevalence within each profile. If multiple profiles share the same leading region, they are further ordered by descending prevalence within that region. Vertical dashed lines separate profiles by the leading region. A map of the regions is provided for reference. In all panels, each bar represents the prevalence of a profile within a specific group. For each bar, statistical significance was assessed using pairwise Z-tests with Bonferroni correction, comparing it to the other groups within the same profile. The largest (i.e., least significant) p-value among the comparisons is shown. Asterisks indicate significance levels (\**p*-value *<* 0.05, \*\**p*-value *<* 0.01, \*\*\**p*-value *<* 0.001). Error bars for 95% confidence intervals are not shown, as the large sample size yields very narrow intervals with minimal impact on interpretability. IMD: Index of Multiple Deprivation.

Socioeconomic deprivation, as measured by quintiles of the IMD, was associated with distinct prevalence patterns across profiles (Fig. 5a,b). Profiles that largely occurred before age 65 (M1–M8, F1–F9, F11) were significantly more prevalent in the most deprived quintile (IMD 5) compared to the least deprived (IMD 1). In contrast, profiles that arose or peaked later in life (M11–M14, M17–M21, F12–F14, F16–F18) were more common among the least deprived.

Pronounced ethnic differences in profile prevalence were also observed (Fig. 5c,d). Among White individuals, mental health-predominant profiles were consistently more prevalent than in other ethnic groups, particularly M1 and F1 (both Anxiety + Depression), and M13 and F14 (both Mental + Physical Long-term Conditions), with prevalence exceeding that of other groups by more than 5%. COPD-related profiles, including M19 (COPD-predominant Respiratory + Cardiovascular) and M16 (Cancer + COPD + Cardiovascular) were also more prevalent among White males. In females, F17 (Hypertension + Cancer + Renal) had a prevalence of 14.58% (95% CI: 14.52–14.64%) among White individuals, more than 5% higher than in other ethnic groups. In comparison, profiles characterised by physical conditions were more prevalent among Black and South Asian populations than in White individuals. Among Black individuals, cardiometabolic profiles such as M10 (Hypertension + Diabetes) and M7 (Hypertension + Diabetes + Mental), along with M17 and F10 (both Cardiometabolic + Renal) showed the highest prevalence, followed by South Asian individuals, with a considerably lower prevalence in White individuals. Among South Asian individuals, profiles characterised by CHD (M11, F11) and those combining asthma and hypertension (M5, M9, F9, F12) were more prevalent than in other ethnic groups. Notably, South Asian males had the highest prevalence of M11 at 24.11% (95% CI: 23.75–24.48%), exceeding all other groups by more than 5%.

Several regional trends were evident (Fig. 5e,f). London was the most ethnically diverse region, with more than 45% of residents identified as non-White, including 14.5% South Asian and 13.5% Black individuals^46^. It had the highest prevalence of profiles (M4, M5, M7, M10, F4, F6, F10) commonly observed in Black and South Asian populations (Fig. 5c,d), reflecting an overlap between ethnic and regional distributions. M10 and F10, the most prevalent profiles among Black and South Asian individuals, showed the highest regional prevalence in the West Midlands, the second most ethnically diverse region in England (11.3% Asian and 4.5% Black)^46^. The regional variation in socioeconomic deprivation was also reflected in the profile distributions. Except for London mainly driven by ethnic diversity, the profiles most prevalent in the least deprived regions of the East of England and the South East (M9, M12, M18–M21, F12, F17, F18) were also the most common in the least deprived IMD quintile (IMD 1; Fig. 5a,b). In contrast, in the North East, the most deprived region in England^47^, seven of the 10 profiles with the highest regional prevalence mirrored those most prevalent in the most deprived IMD quintile (IMD 5).

### Biological factors with multimorbidity

We next examined the multimorbidity profiles in association with 45 routinely measured clinical markers (Fig. 6). Relevance was defined as the extent to which abnormal marker values (elevated or reduced) contributed to the correct classification of each profile (Methods).

**Figure 6:**
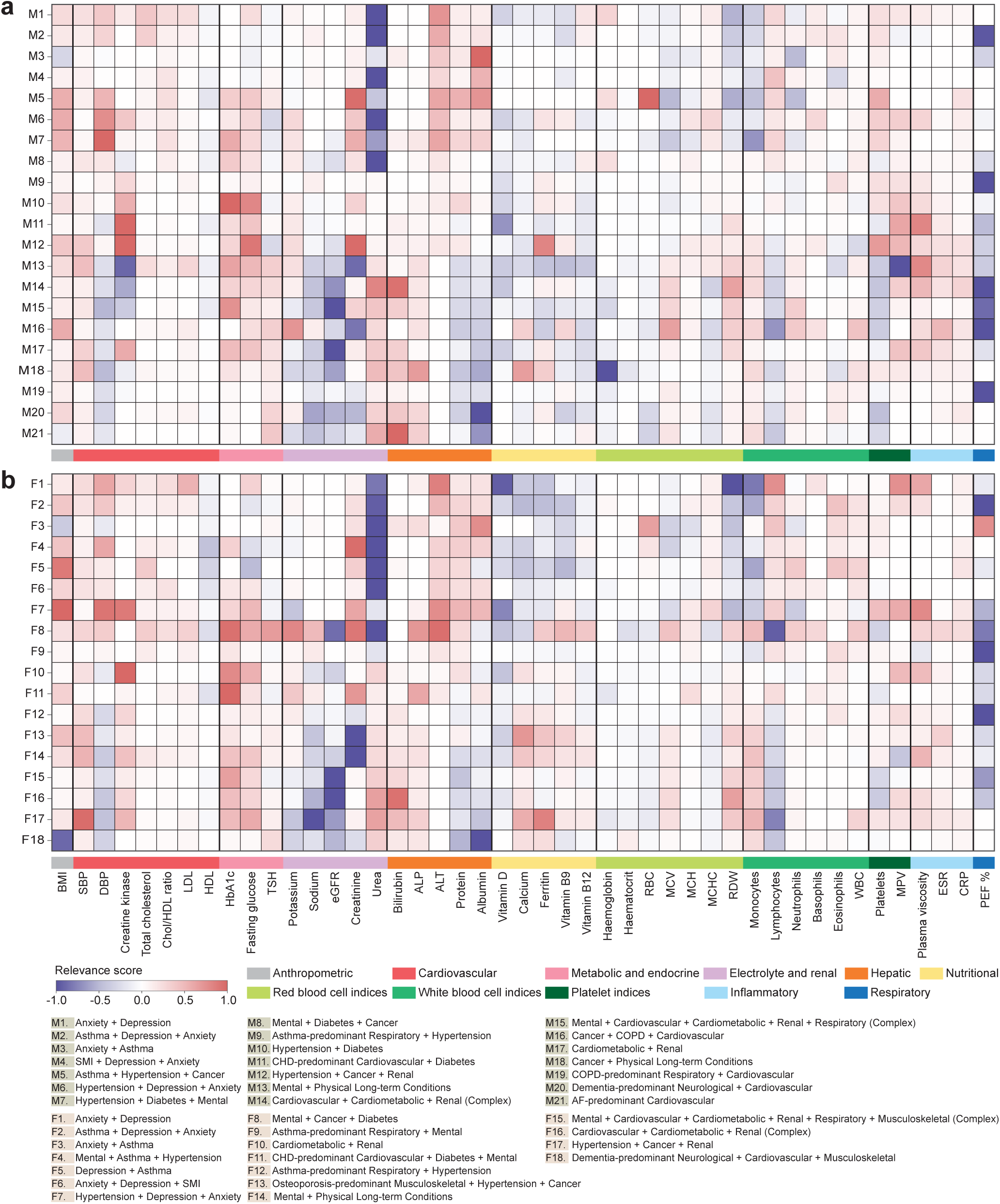
Relevance of clinical markers across multimorbidity profiles for a, Males and b, Females. The heatmaps show the clinical relevance of each marker (columns) to each multimorbidity profile (rows), as determined by an interpretable machine learning framework (Methods). The hue (red/purple) indicates whether the marker value is abnormally high or low relative to a reference range, and the intensity (gradient) reflects the strength of its association. Markers are grouped by clinical system or physiological process (Supplementary Table S2). Beeswarm plots with SHAP values for each profile are provided in Supplementary Fig. S11–S12.

Across nearly all profiles, elevated BMI and systolic blood pressure (SBP) showed high relevance. In contrast, diastolic blood pressure (DBP) followed an age-dependent pattern: elevated DBP characterised profiles occurring under age 65, whereas reduced DBP predominated in cardiovascular profiles among those aged 65 years and older.

Respiratory-related profiles (M2, M3, M5, M9, M15, M16, M19, F2–F5, F9, F12, F15) were distinguished by reduced peak expiratory flow percentage (PEF %) (Fig. S9a). Neurological profiles (M20, F18) exhibited the strongest relevance for reduced albumin, and F18 also showed a strong relevance for reduced BMI (Fig. S9b). Cancer-related profiles (M5, M8, M12, M16, M18, F8, F13, F17) were associated with elevated inflammatory markers including erythrocyte sedimentation rate (ESR), C-reactive protein (CRP) and plasma viscosity (Fig. S9c). The musculoskeletal profile F13 was marked by elevated calcium and reduced vitamin D. Early-life mental health profiles were associated with reduced urea and elevated hepatic markers including alanine aminotransferase (ALT), total protein and albumin. Within the cardiometabolic-related profiles, those with high diabetes prevalence (M7, M8, M10, M11, M14, M15, M17, F8, F10, F11, F15, F16) demonstrated strong associations with markers of glucose metabolism, particularly HbA1c and fasting glucose (Fig. S10a). Cardiovascular profiles characterised by heart-failure prevalence (M14, M21, F16) showed relevance for elevated bilirubin. Renal-related profiles (M12, M14, M15, M17, F10, F15–F17) were defined by markers of renal dysfunction, including reduced estimated glomerular filtration rate (eGFR), along with elevated urea and reduced sodium. Finally, the four complex profiles (M14, M15, F15, F16) demonstrated multisystem involvement through associations with diverse markers, including eGFR, PEF%, HbA1c and fasting glucose.

## Discussion

Multimorbidity presents a critical challenge to both individual health and healthcare systems. In this large, population-based longitudinal study of 3.3 million individuals in England, we present the most comprehensive characterisation of multimorbidity to date. Using EHRs that span the life course, this study is, to our knowledge, the first to identify distinct multimorbidity profiles, their life-course trajectories, associated health burdens, and underlying social and biological factors at a national level. Our findings reveal clinically plausible profiles that evolve with age, assess the differential burden of multimorbidity on mortality and hospitalisation, highlight inequalities shaped by social factors, and suggest patterns in clinical markers associated with specific profiles. This new way of characterising multimorbidity can provide valuable insights for clinical practice and public health policy.

Several findings stand out. First, our analysis reveals the long persistence of specific multimorbidity profiles throughout the life course, particularly the mental health-only profile during early and middle adulthood, and cardiometabolic and cardiovascular profiles emerging in middle adulthood and persisting into older age. These patterns highlight the long-lasting burden of mental health conditions and the cumulative impact of metabolic and vascular diseases. Clear sex-specific patterns were also evident in our study. Males were more frequently represented in cardiovascular, respiratory, and cancer-related profiles, while females were more commonly represented in mental, neurological, and musculoskeletal profiles. These differences reflect a combination of behavioural and biological factors: higher rates of smoking and alcohol consumption among males may contribute to COPD and cancer risk, while oestrogen confers cardiovascular protection for females until menopause. Differences in stress exposure, help-seeking and health service variation may also play a role. For example, females are more likely to seek care for depression and present with symptoms that align more closely with clinical diagnostic criteria, while males with comparable scores on standardised mental health assessments are less frequently diagnosed. Our findings highlight the need for prevention strategies tailored to sex-specific risks, including reducing cardiovascular risk in males and improving mental health support for females.

In addition, our study makes an important novel contribution to the literature by mapping multimorbidity trajectories to show how different progression pathways evolve throughout the life course. For example, we identified transitions from a healthy state to cardiometabolic conditions (e.g., hypertension and diabetes) and subsequently to profiles combining cardiometabolic and renal disorders. We also observed transitions from asthma-predominant profiles to COPD-predominant profiles, likely driven by chronic inflammation of the airways and environmental exposures such as smoking and air pollution. Similarly, progression from cardiometabolic-predominant profiles to more complex profiles incorporating cardiovascular conditions may reflect cumulative endothelial dysfunction, atherosclerotic burden, and lifestyle factors such as physical inactivity. These life-course pathways expand previous longitudinal observations, and reveal clearly defined critical transition points and age windows, which could serve as valuable opportunities for preventive interventions.

Interestingly, approximately half of individuals with multimorbidity remained within a single profile over the life course, while the remainder transitioned between multiple distinct profiles. These observations suggest that tailored preventive strategies may be needed: those with stable multimorbidity may benefit from targeted management of specific conditions, whereas those following dynamic trajectories may require broader strategies to interrupt progression to more complex multimorbidity.

Our findings also shed light on how specific conditions shape the burden of multimorbidity. Profiles exhibiting high-risk physical conditions, particularly cardiovascular and renal conditions, were associated with the highest mortality, in line with previous studies^18,48^. In contrast, highly complex profiles encompassing multiple systems associated with elevated hospitalisation rates, despite relatively lower mortality, reflect the resource-intensive demands of complex multimorbidity management. Neurological profiles exhibited particularly high mortality but only moderate hospitalisation, possibly due to management in long-term care facilities or community settings, limited therapeutic options, or transitions to palliative care in advanced stages of the disease. These findings emphasise the importance of moving beyond simple condition counts in the multimorbidity assessment and considering disease interactions and clinical severity for outcome management and healthcare resource allocation.

The study further highlights pronounced disparities in multimorbidity, shaped by socioeconomic deprivation, ethnicity, and geography. Individuals from deprived backgrounds experienced earlier onset and greater burden of multimorbidity before age 65, whereas later-life multimorbidity was more prevalent among less deprived groups. This probably reflects the interplay of survival bias, differential access to healthcare, and the protective effects of socioeconomic advantage: deprived populations experience earlier disease onset and premature mortality, whereas more affluent individuals are more likely to survive into older age, where the cumulative nature of ageing leads to higher prevalence of complex multimorbidity profiles. In addition to socioeconomic deprivation, ethnicity played a significant role. White populations showed a higher prevalence of mental health-predominant profiles, potentially reflecting cultural and systemic determinants in awareness, help-seeking, and diagnosis. At the same time, minority ethnic populations may face reduced access to mental health services. In addition, White populations were more represented in cancer-related profiles, likely due to greater participation in screening programmes, which in turn increases the incidence rates of certain cancers. This is also associated with a higher prevalence of risk factors, such as smoking and obesity, compared to other ethnic groups. In contrast, South Asian and Black populations showed a higher prevalence in cardiovascular and cardiometabolic profiles, which is consistent with differences in insulin resistance and dietary patterns, including significantly higher salt and carbohydrate consumption^49^. These findings confirm the role of social factors, including structural healthcare inequalities, in shaping multimorbidity, highlighting the need for targeted interventions, including optimised cardiometabolic management, equitable screening programmes, and enhanced mental health outreach for at-risk populations.

Regional variations further reflected the interplay of ethnicity and socioeconomic deprivation in multimorbidity. Ethnic diversity emerged as the dominant factor in London, while socioeconomic deprivation had a stronger influence in the North East. Elsewhere, the relationship between these factors and multimorbidity was more heterogeneous, highlighting the importance of examining social factors on granular geographical scales where population characteristics are more homogeneous. We acknowledge, however, that these disparities arise not only from the investigated social factors but also from systemic biases in diagnosis, unequal environmental exposures, historical mistrust of healthcare systems and other structural determinants, necessitating interventions that address both clinical management and the broader contextual forces shaping health.

An important novelty of our study lies in the systematic integration of clinical markers with multimorbidity profiles through an interpretable machine learning framework. Several observed associations, such as elevated BMI and SBP across all profiles, are consistent with established roles of obesity and hypertension in multimorbidity development^50^, while the associations of calcium and vitamin D with the musculoskeletal profile align with known metabolic imbalances underlying osteoporosis^51^. The consistency of these associations across a large, representative population supports the external validity of the framework and highlights its utility in identifying clinically relevant patterns. Importantly, this study extends beyond prior studies in several key aspects. First, unlike most efforts that focus on single conditions, our framework enables fine-grained, profile-specific marker association analysis across multimorbidity profiles. Second, the model is trained on a uniquely large and diverse primary care dataset, providing greater power and population generalisability compared to earlier studies typically based on smaller or more selective cohorts.

Importantly, we identified several underexplored or novel marker associations with multimorbidity profiles. For example, the age-dependent pattern with DBP reflects the progression toward isolated systolic hypertension in ageing populations, mediated by progressive arterial stiffening and loss of vascular compliance^52^. The elevation of hepatic markers in mental health-predominant profiles may reflect unmeasured confounding, such as alcohol use, psychotropic medication effects, or systemic inflammation, and suggest biological pathways not previously recognised in this context. Similarly, the prominence of albumin in neurological profiles and bilirubin in heart failure profiles provides interpretable markers, plausibly linked to cognitive decline and hepatic congestion, respectively. Together, these findings underscore the translational potential of our approach. By linking clinical markers to multimorbidity profiles, our framework enables more precise risk stratification, earlier detection of individuals at elevated multimorbidity risk, and the development of tailored screening and management protocols. Its adaptability supports further applications across diverse populations and broader marker panels to enhance diagnostic decision-making and inform therapeutic strategies.

Our study design should be interpreted in light of several limitations. First, we assume that all 18 conditions persist after diagnosis, potentially overlooking recovery, undiagnosed or unrecorded underlying conditions, and misrepresenting acute events such as stroke, which have a transient onset but lasting impact. Second, the use of CPRD data introduces inherent biases, particularly related to access to primary care, influenced by factors such as the inverse care law, where individuals with higher needs may receive inadequate care. In addition, the quality of general practitioner coding, which is heavily influenced by the Quality and Outcomes Framework (QOF)^53^, could affect the accuracy of diagnoses and clinical marker measurements, although the large sample size and advanced analytical methods help mitigate these concerns. Third, the design of this observational study inherently restricts causal inferences, especially in analyses involving social factors and associations of clinical markers. Furthermore, while we examined the influence of social factors on multimorbidity, we did not account for their potential interactions or confounding effects, which may overlook the complex interplay among these factors in shaping multimorbidity. Lastly, we were unable to fully adjust for the effects of medications that may influence clinical marker levels, affecting the clinical interpretation of the identified markers. These limitations highlight important avenues for future research while not diminishing the value of our population-level findings.

In conclusion, our work provides a comprehensive characterisation of multimorbidity throughout the life course in a nationally representative population, revealing the complex and dynamic nature of multimorbidity, shaped by disease interactions, distinct progression pathways, differential burden patterns, and social and biological factors. Collectively, these findings highlight the need for adaptive and risk-stratified public health frameworks to inform targeted prevention strategies, optimise healthcare resource allocation, and guide evidence-based policies to address the growing challenge of multimorbidity.

## Data Availability

The data underlying this article is provided by the UK CPRD electronic health record database, which is only accessible to researchers with protocols approved by CPRD Research Data Governance.

## Methods

### Study population

This work was conducted as part of CoMPuTE project (Complex Multiple Long Term Conditions—Phenotypes, Trends, and Endpoints). We conducted a longitudinal cohort study using anonymised EHRs from CPRD^34^, which received ethical approval (CPRD reference: 22 001771). Our cohort consisted of 6,671,245 individuals, including 3,314,652 who developed multimorbidity (≥2 of 18 selected chronic conditions), 1,226,245 who remained condition-free, and 2,130,348 with a single condition at the end of study. Participants were followed from birth until the first occurrence of death or study exit no later than 31 December 2019. Study exit was defined as the earliest transfer out or the last practice download date.

### Chronic conditions

We studied 18 chronic conditions selected based on prevalence and clinical significance across major disease categories: mental health (anxiety, depression, serious mental illnesses), respiratory (asthma, chronic obstructive pulmonary disease), metabolic (diabetes), cardiovascular (hypertension, coronary heart disease, stroke or transient ischaemic attack, atrial fibrillation, heart failure, peripheral arterial disease), renal (chronic kidney disease), neurological (dementia, Parkinson’s disease), musculoskeletal (osteoporosis, rheumatoid arthritis), and oncological (cancers excluding non-melanoma skin cancers). Of these, 16 conditions were selected from the QOF indicators, with anxiety and Parkinson’s disease additionally included due to their increasing prevalence and clinical relevance with age. SNOMED CT code lists (https://github.com/ndpchs-cprd/CPRD-22-001771-CoMPuTe/tree/main/Codelists) were used to ensure a consistent identification of conditions. The first recorded diagnosis of each condition was assumed to reflect its presence thereafter. The selected conditions collectively represent the majority of the multimorbidity impact, enabling examination of both physical-mental health interactions and cross-system disease clustering patterns.

### Other measures and variables

We included CPRD data on sex, date of birth, dates of diagnosis for each condition, IMD quintiles (as a measure of socioeconomic deprivation), ethnicity, and geographic region based on the location of the registered general practitioner. The characteristics of the primary study cohort by social factors are summarised in Supplementary Table S1. Hospitalisation data were obtained through linkage with HES^42^, and mortality data were linked from ONS^41^. In addition, we retrieved 45 conventional clinical markers from CPRD records (Supplementary Table S2).

### Multimorbidity profile identification

For each individual, we map the diagnosis of each condition to nine age bands spanning the life course (from *<*18 to ≥85 years), creating 18-dimensional binary health state vectors indicating which specific conditions had been diagnosed by the end of each age band. Within each age-sex stratum, we applied LCA^17^ to individuals with multimorbidity (≥2 conditions) to identify distinct clusters. Following previous studies^7,18^, the optimal number of clusters was determined on the basis of model parsimony using the Bayesian information criterion (BIC), Akaike information criterion (AIC) and consistent AIC (cAIC), which balance goodness of fit against model complexity to minimise overfitting. Final selection further incorporated clinical relevance and interpretability, as established through successive rounds of expert-panel review and consensus meetings within the CoMPuTE consortium. For each identified cluster, we computed two key metrics: condition prevalence, defined as the proportion of individuals within a cluster having each condition; and condition exclusivity, defined as the proportion of individuals with a specific condition in a given age band who belonged to that particular cluster (Supplementary Fig. S2–S3). Subsequently we applied agglomerative hierarchical clustering using Ward’s method to the vectors of condition prevalence and exclusivity, thereby quantifying cluster similarity across age bands (Supplementary Fig. S4). Clusters within each sex group were merged based on thresholds informed by clinical interpretability and hierarchical clustering results (Supplementary Note S1.1), thus forming multimorbidity profiles. Each multimorbidity profile was characterised by its distinct composition of chronic conditions (Supplementary Fig. S5) and named according to a convention detailed in Supplementary Note S1.2. This approach ensured accurate identification of sex- and age-specific profiles while capturing multimorbidity patterns consistently across different strata. LCA was implemented using the StepMix package^54^, and hierarchical clustering was performed using the SciPy package^55^ in Python v3.8.

### Multimorbidity trajectory reconstruction

Using the multimorbidity profiles identified for each individual across the age bands, we identified the longitudinal trajectories of multimorbidity. We also identified states in which individuals were free of all 18 chronic conditions (denoted “H”), had a single condition (“S”), had died (“D”), or had exited the study (“E”).

### Multimorbidity burden assessment

We assessed the burden of multimorbidity using three metrics: mortality, hospitalisation rate, and hospitalisation prevalence. Mortality for each multimorbidity profile was calculated within each age band as the proportion of individuals who died while in that profile relative to the total number of individuals who experienced the profile in that age band. The hospitalisation rate was defined as the mean number of hospitalisations per year among individuals within a specific profile and age band. The prevalence of hospitalisation represented the proportion of individuals hospitalised at least once during the age band relative to the total number of people who experienced the profile in that age band.

### Statistical analysis with social factors

We analysed the association between the multimorbidity profiles and key social factors: socioeconomic deprivation, ethnicity, and geographic region. For each profile, we calculated its prevalence across subgroups within each social factor and performed statistical significance tests using pairwise Z-tests with Bonferroni correction. For each subgroup-profile combination, we compared prevalence with all other subgroups within the same profile and reported the least significant p-value from these comparisons.

### Interpretable machine learning framework with biological factors

Although clinical markers are well-established indicators for individual conditions, their role in multimorbidity remains poorly characterised^6,10^. The synergistic effects of coexisting conditions can substantially alter clinical marker patterns, rendering condition-level associations insufficient for identifying profile-level markers. To address this, we developed an interpretable machine learning framework using XGBoost^43^ combined with a novel reference-adjusted SHAP^44,45^ to quantify the relevance of clinical markers to specific multimorbidity profiles in a data-driven manner. This framework offers several advantages: XGBoost inherently handles missing values and maintains scale invariance across heterogeneous marker distributions, while the reference-adjusted SHAP quantifies individual marker contributions relative to clinical reference ranges, thereby improving clinical interpretability.

For each individual in each age band, we extracted the most recent measurement of the 45 clinical markers, aligning with the latest diagnosis information used for profile identification. This resulted in clinical marker vectors that represented individuals’ biological states in specific age bands. Missing marker values were explicitly indicated in vectors, and each clinical marker vector was labelled according to the individual’s multimorbidity profile in the corresponding age band. We then sampled 10,000 clinical marker vectors per profile for each sex, creating two datasets comprising 210,000 male and 180,000 female samples. These datasets were divided into training subsets (80%) and testing subsets (20%). An XGBoost classifier was trained to predict multimorbidity profiles based on clinical markers, formulated as a multiclass classification task. Owing to XGBoost’s inherent ability to handle missing values and scale invariance, we did not perform additional preprocessing or normalisation. The key hyperparameters included a learning rate of 0.1, a maximum tree depth of 12, and 500 boosting iterations. The performance metrics of the model demonstrated robust discrimination, with males achieving an area under the receiver operating characteristic curve (AUROC) of 0.894, area under the precision-recall curve (AUPRC) of 0.384, and top-k accuracy of 0.383 (k=1), 0.677 (k=3), and 0.814 (k=5). Female models showed comparable performance (AUROC=0.891, AUPRC=0.413) with top-1 (0.402), top-3 (0.703) and top-5 (0.841) accuracy. In practice, a top-k prediction of the most probable profiles could enable clinicians to prioritise preventive and monitoring strategies tailored to the likeliest progression pathways. To improve clinical interpretability, we implemented a reference-adjusted SHAP methodology. Traditional SHAP analysis can highlight markers that are valuable for classification but clinically irrelevant, such as markers of the normal range incorrectly identified as important indicators. Our approach focused specifically on markers whose abnormal values (outside the clinically defined reference ranges) positively contributed to the correct identification of multimorbidity profiles. We defined clinical relevance as the mean SHAP value among samples with positive SHAP values and clinical marker values beyond their normal reference ranges (Fig. S11–S12). This refined analysis identified clinical markers that are associated with multimorbidity profiles, potentially serving as early clinical indicators for these profiles. We conducted these analyses using the xgboost package^43^ and the SHAP package^45^ in Python v3.8.

## Ethical approval

The CPRD database is approved by the East Midlands–Derby Research Ethics Committee, reference 05/MRE04/87 for public health research according to approved research protocols, and the research protocol for this study received specific approval from the CPRD’s Research Data Governance (reference 22 00177).

## Data availability

Data supporting the findings of this study are available in the article and its Supplementary information. The data underlying this article is provided by the UK CPRD electronic health record database, which is only accessible to researchers with protocols approved by CPRD’s Research Data Governance.

## Code availability

The code used for this study is available at https://github.com/liuyuaa/ CoMPuTE-Multimorbidity-in-England.

## Acknowledgements.

This project is funded by the NIHR Programme Grants for Applied Research Programme (NIHR204406). The views expressed are those of the author(s) and not necessarily those of the NIHR or the Department of Health and Social Care. This study is based in part on data from the Clinical Practice Research Datalink obtained under licence from the UK Medicines and Healthcare products Regulatory Agency. The data is provided by patients and collected by the NHS as part of their care and support. The interpretation and conclusions contained in this study are those of the authors alone. T. Zhu was supported by the Royal Academy of Engineering under the Research Fellowship scheme. We thank X. Li for clinical advice and Y. Jiang for visualisation advice.

## Authors contributions

Y.L. and T.Z. conceived and designed the study with advice from all authors. C.B., C. W. D. and S.S. performed data acquisition and curation. Y.L. developed the analytical framework, implemented machine learning methods, conducted all analyses, and drafted the manuscript. Y.L., C.B., C.W.D., C.P., D.G., R.P. and T.Z. led the initial interpretation of results. All authors discussed the findings, contributed to manuscript revisions, and approved the final version.

## Consortia

### CoMPuTE Consortium

Sami Adnan^2^, Henrique Aguiar^1^, Anica Alvarez Nishio^2^, Amitava Banerjee^4^, Clare Bankhead^2^, Derrick Bennett^5^, Kamaldeep Bhui^2,6^, Benjamin Cairns^7^, Mei Chan^5^, Andrew Clegg^8^, Caroline Cupit^2^, Ashkan Dashtban^4^, Cynthia Wright Drakesmith^2^, Margaret Glogowska^2^, David Gonzalez^3^, Mackenzie Graham^5^, Odessa Hamilton^2^, Ishbel Henderson^2^, Carl Heneghan^2^, Joanna Lach^2^, Mary Logan^2^, Sapfo Lignou^5^, Yu Liu^1^, Micheal McKenna^2^, Brian Nicholson^2^, Igho Onakpoya^2^, Rafael Perera-Salazar^2^, Nicola Pidduck^2^, Catherine Pope^2^, John Powell^2^, Samuel Relton^8^, Eve Rodgers^6^, Anna Seeley^2^, Mark Sheehan^5^, James Sheppard^2^, David Steinsaltz^9^, Subhashisa Swain^2^, Tazeen Tahsina^2^, Paul Taylor^4^, Oliver Todd^8^, Apostolos Tsiachristas^2^, Anthony Webster^9^, Tingting Zhu^1^

^1^Department of Engineering Science, University of Oxford, Oxford, UK.

^2^Nuffield Department of Primary Care Health Sciences, University of Oxford, Oxford, UK.

^3^Discipline of General Practice, University of Adelaide, Adelaide, Australia

^4^Institute of Health Informatics, University College London, London, UK.

^5^Nuffield Department of Population Health, University of Oxford, Oxford, UK.

^6^Department of Psychiatry, University of Oxford, Oxford, UK.

^7^Our Future Health, London, UK.

^8^School of Medicine, University of Leeds, Leeds, UK.

^9^Department of Statistics, University of Oxford, Oxford, UK.

