## Supplementary material for "Identifying profiles, trajectories, burden, social and biological factors in 3.3 million individuals with multimorbidity in England": s

### Supplementary Notes

### S1 Note 1: Identification of multimorbidity profiles

#### S1.1 Threshold selection for merging clusters

To merge identified clusters across different age bands into unified multimorbidity profiles, we determined thresholds for hierarchical clustering using the following criteria:

- Clusters from the same age band were not merged.
- Only clusters from adjacent age bands were eligible for merging, ensuring multimorbidity profiles do not span non-consecutive age groups.

Using these criteria, we identified the merging threshold as the smallest hierarchical clustering distance that met both conditions.

#### S1.2 Multimorbidity profile naming convention

When naming multimorbidity profiles, we consider several key clinically meaningful factors:

- **Common conditions reflecting population characteristics.** Highly prevalent conditions such as depression or hypertension, though often not profile-specific, characterise substantial proportions of cluster populations and thus are relevant for clinical context.
- **Clinically significant rare conditions.** Conditions with a lower prevalence within a profile, but significant clinical implications or distinctive diagnostic value, were considered as important differentiators.
- **Within-age-band differentiation.** Conditions or body systems uniquely prevalent within certain profiles relative to others of the same age group were emphasised, clarifying

between-profile distinctions and improving clinical interpretability.

Consequently, we defined the following guidelines for the naming of multimorbidity profiles:

- **Conditions with 100% prevalence.** Conditions universally present within a profile were always included in the profile name (e.g., Anxiety or Depression in names of M1, M3, F1, F3, and F5).
- **Complex profiles.** Profiles characterised by an average of more than four conditions per individual were labelled with the suffix "Complex".
- **Condition inclusion criteria.**
    - Profile names included up to three specific conditions or body systems, except for profiles designated as "Complex".
    - Conditions included in names must meet at least one of the following criteria:
        - Within-profile prevalence greater than 40% (e.g., conditions included in names of M10 and F2), or
        - Substantially higher within-profile prevalence relative to other profiles in the same age band (e.g., Cancer in names of M5 and F13, Cancer and Diabetes in the name of M8).
- **Use of system-level naming.**
    - When multiple conditions from the same physiological system were present, the system name was used (e.g., Cardiovascular, Cardiometabolic, Renal, Respiratory, and Musculoskeletal systems in the name of F15).
    - If one condition within a system demonstrated a significantly higher prevalence compared to others, the system was explicitly labelled as "xx-predominant" (e.g., Osteoporosis-predominant in the name of F13).
- **Ordering of conditions and systems.** Conditions and systems were listed in descending order of prevalence within the profile.

### Supplementary Figures

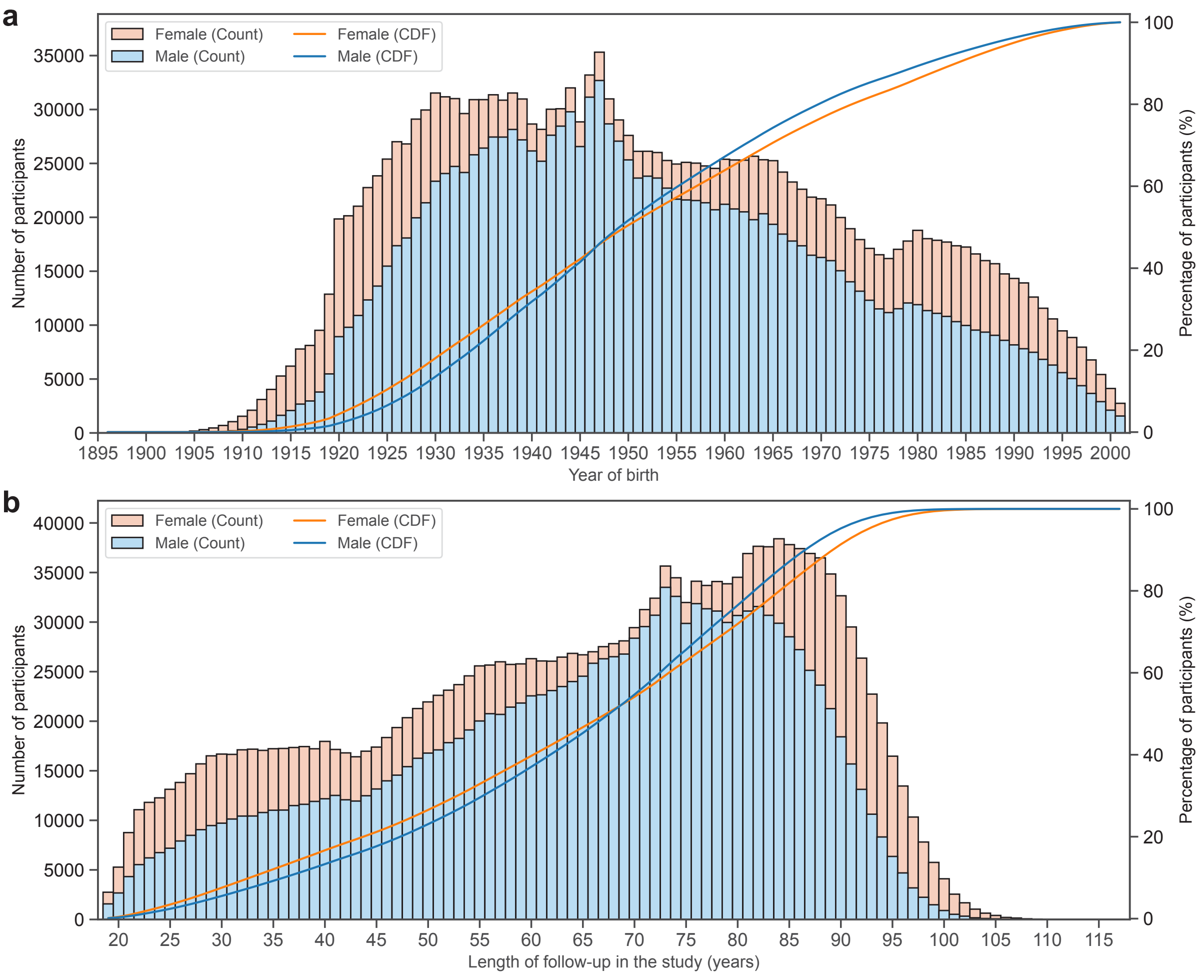

**Figure S1: Participant birth year and follow-up duration. a,** Distribution of participants by year of birth and sex. **b,** Length of follow-up in the study (years), stratified by sex. The stacked bars represent the absolute counts of participants, while the lines indicate the cumulative percentages.

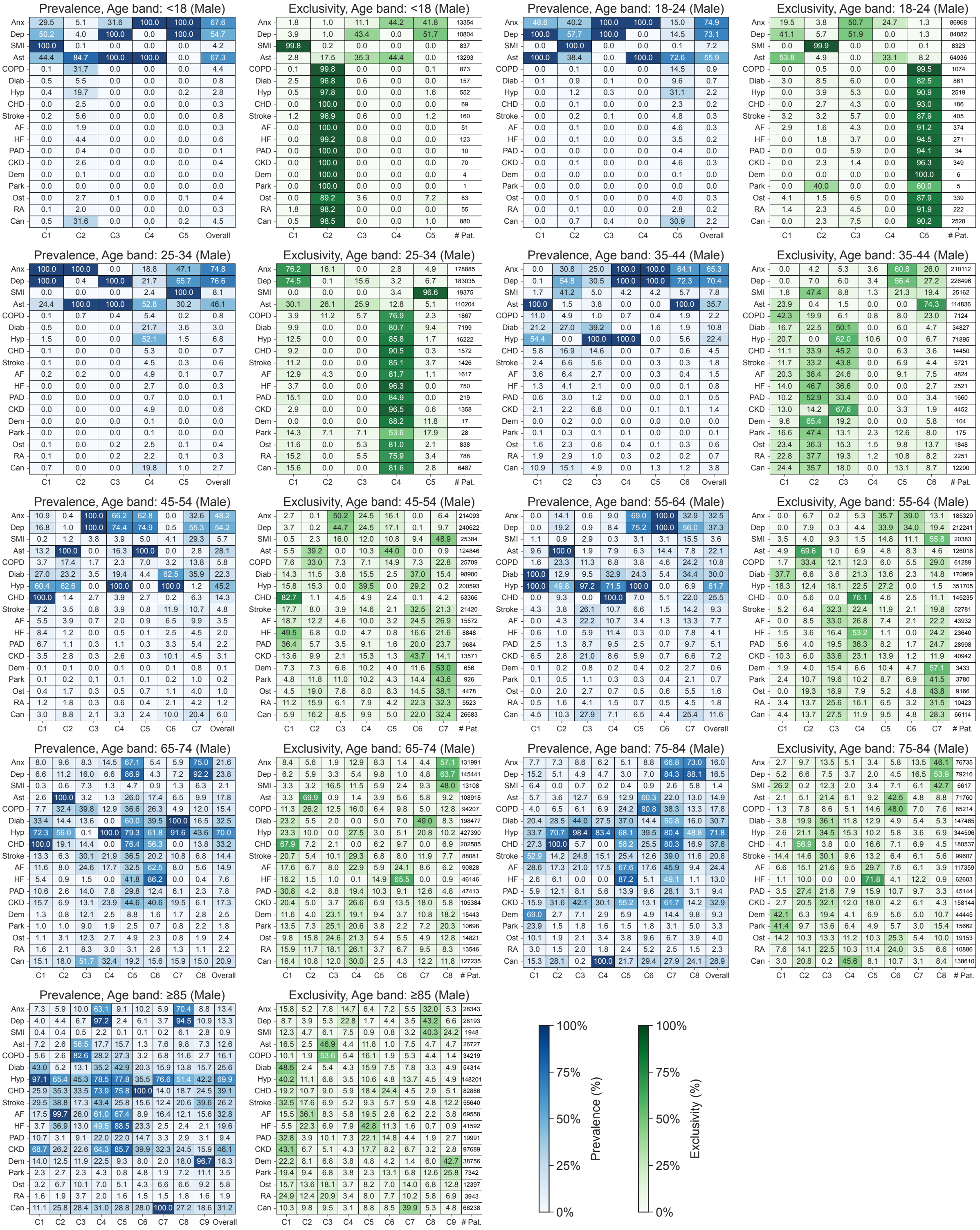

**Figure S2: Condition prevalence and exclusivity for clusters in different age bands for males.** Condition abbreviations: anxiety (Anx), depression (Dep), serious mental illness (SMI), asthma (Ast), chronic obstructive pulmonary disease (COPD), diabetes (Diab), hypertension (Hyp), coronary heart disease (CHD), stroke or transient ischaemic attack (Stroke), atrial fibrillation (AF), heart failure (HF), peripheral arterial disease (PAD), chronic kidney disease (CKD), osteoporosis (Ost), rheumatoid arthritis (RA), cancer excluding non-melanoma skin cancers (Can).

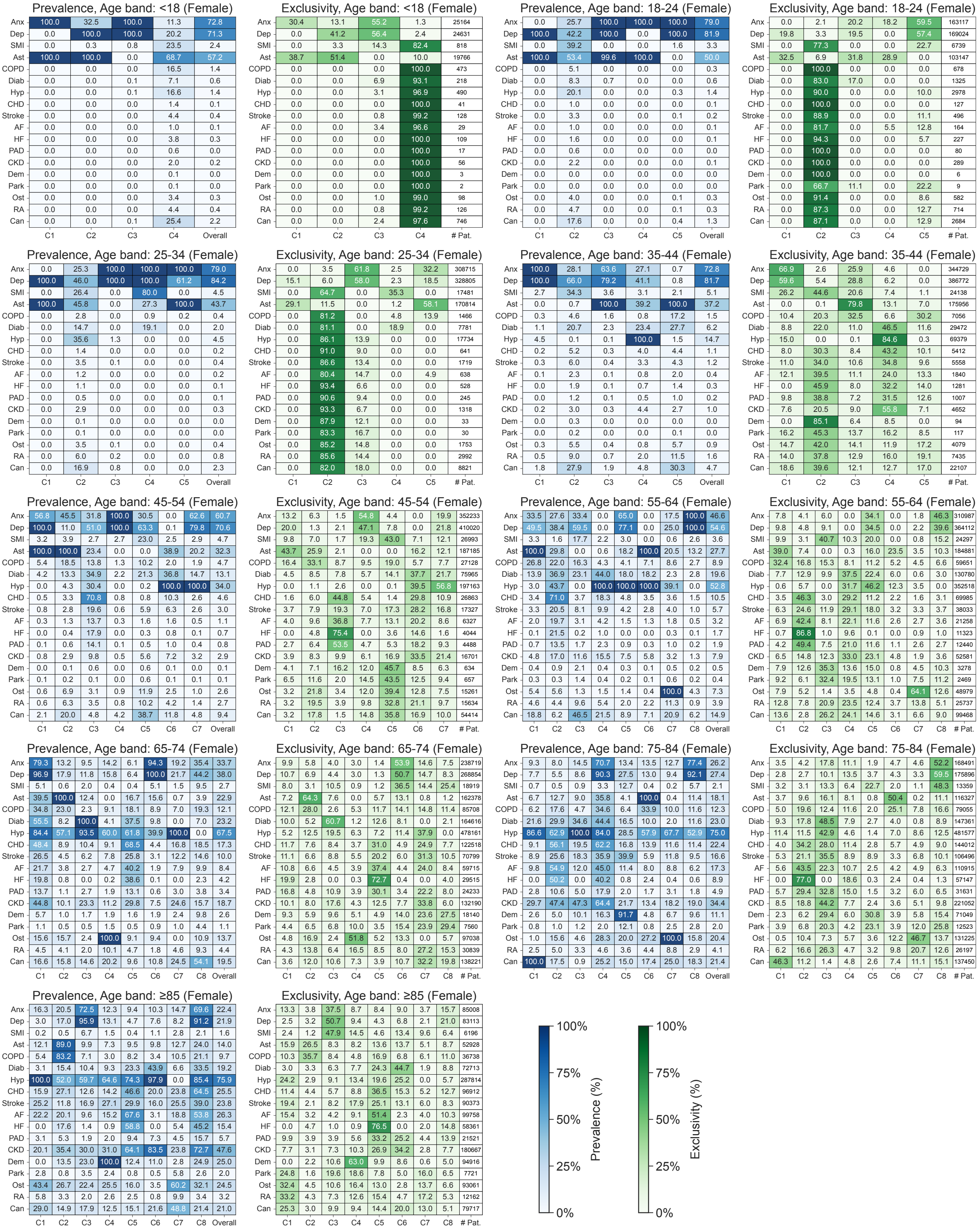

**Figure S3: Condition prevalence and exclusivity for clusters in different age bands for females.** Condition abbreviations: anxiety (Anx), depression (Dep), serious mental illness (SMI), asthma (Ast), chronic obstructive pulmonary disease (COPD), diabetes (Diab), hypertension (Hyp), coronary heart disease (CHD), stroke or transient ischaemic attack (Stroke), atrial fibrillation (AF), heart failure (HF), peripheral arterial disease (PAD), chronic kidney disease (CKD), osteoporosis (Ost), rheumatoid arthritis (RA), cancer excluding non-melanoma skin cancers (Can).

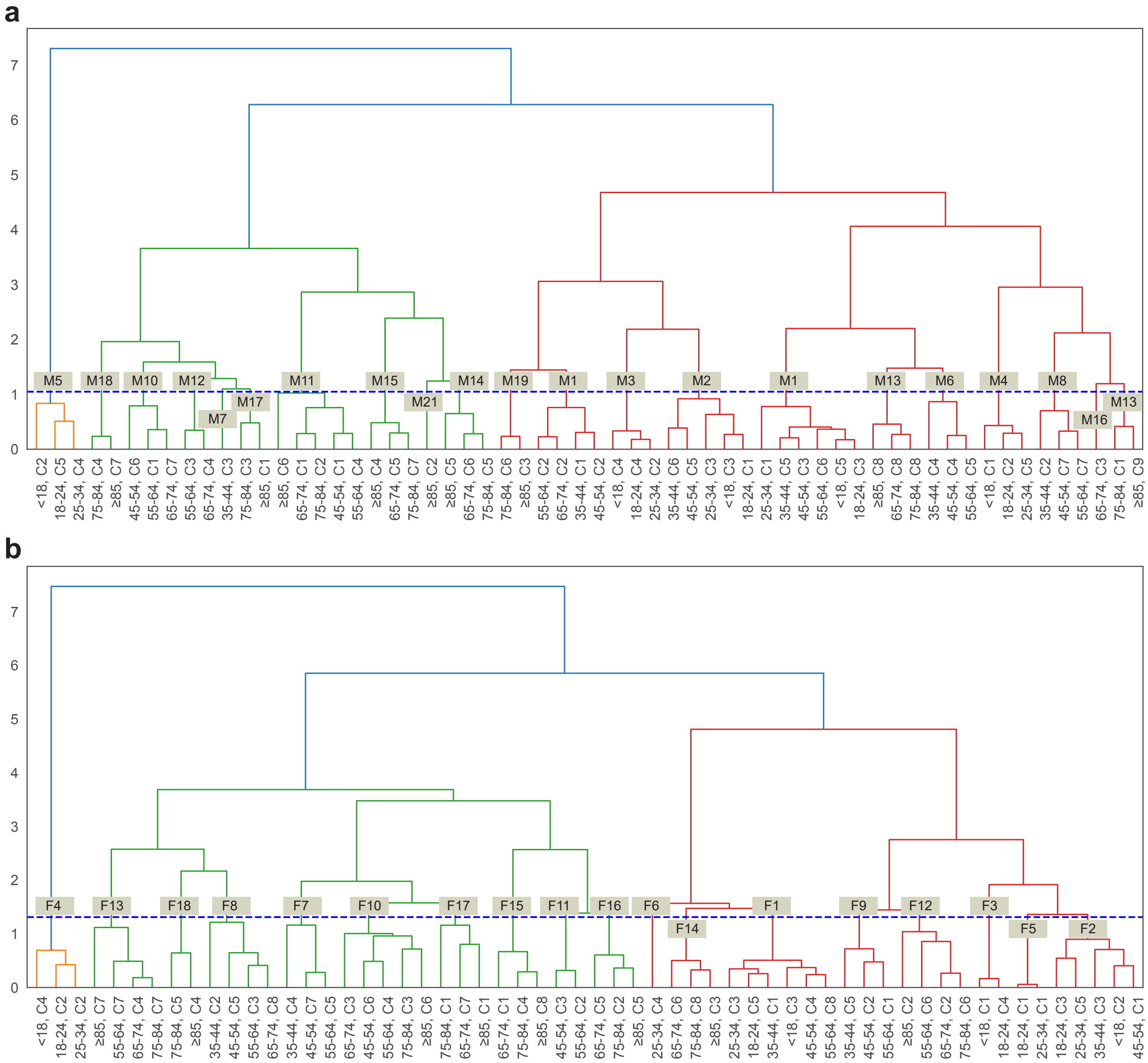

**Figure S4: The cluster merging process for multimorbidity profiles. a,** Clusters merged from different age bands for males. **b,** Clusters merged from different age bands for females. The blue dashed lines indicate the threshold to stop the merging process, determined by the rules in Note S1.1.

**a**

| | Anx | Dep | SMI | Ast | COPD | Diab | Hyp | CHD | Stroke | AF | HF | PAD | CKD | Dem | Park | Ost | RA | Can |
|---|---|---|---|---|---|---|---|---|---|---|---|---|---|---|---|---|---|---|
| M1 | 100.00 | 100.00 | 2.37 | 8.84 | 1.19 | 2.13 | 0.44 | 1.54 | 0.51 | 0.47 | 0.01 | 0.18 | 0.20 | 0.04 | 0.08 | 0.22 | 0.21 | 1.68 |
| M2 | 51.65 | 82.04 | 3.66 | 100.00 | 2.68 | 1.97 | 2.32 | 0.97 | 0.34 | 0.42 | 0.07 | 0.10 | 0.12 | 0.02 | 0.03 | 0.33 | 0.22 | 1.15 |
| M3 | 100.00 | 0.21 | 0.00 | 100.00 | 0.37 | 0.00 | 0.00 | 0.00 | 0.00 | 0.12 | 0.00 | 0.00 | 0.00 | 0.00 | 0.00 | 0.00 | 0.00 | 0.00 |
| M4 | 44.53 | 62.88 | 100.00 | 33.09 | 0.18 | 2.69 | 1.37 | 0.04 | 0.24 | 0.08 | 0.02 | 0.00 | 0.06 | 0.01 | 0.03 | 0.08 | 0.12 | 0.88 |
| M5 | 16.99 | 18.90 | 1.80 | 59.11 | 9.16 | 18.12 | 45.48 | 4.52 | 4.68 | 4.65 | 2.99 | 0.62 | 4.66 | 0.07 | 0.05 | 2.85 | 2.32 | 22.91 |
| M6 | 69.10 | 75.90 | 3.50 | 10.35 | 3.04 | 21.14 | 100.00 | 5.39 | 5.16 | 2.63 | 0.36 | 1.79 | 4.26 | 0.23 | 0.23 | 0.47 | 0.62 | 4.88 |
| M7 | 25.05 | 30.49 | 4.98 | 3.82 | 0.97 | 39.17 | 100.00 | 14.64 | 5.62 | 2.67 | 2.07 | 1.25 | 6.75 | 0.04 | 0.05 | 0.63 | 0.98 | 4.92 |
| M8 | 32.39 | 55.54 | 24.64 | 5.07 | 17.29 | 33.39 | 3.86 | 16.36 | 11.65 | 10.89 | 6.09 | 7.14 | 5.12 | 1.64 | 1.42 | 4.41 | 4.08 | 21.88 |
| M9 | 8.29 | 10.79 | 0.95 | 100.00 | 23.59 | 16.46 | 54.91 | 10.43 | 4.38 | 5.31 | 1.05 | 1.62 | 4.03 | 0.34 | 0.55 | 2.25 | 1.83 | 12.52 |
| M10 | 2.61 | 3.30 | 1.97 | 5.66 | 3.46 | 90.02 | 96.31 | 0.04 | 9.18 | 5.29 | 0.83 | 4.33 | 13.19 | 0.82 | 0.45 | 0.63 | 1.31 | 11.01 |
| M11 | 8.54 | 7.94 | 0.21 | 5.46 | 6.48 | 30.68 | 68.48 | 100.00 | 12.18 | 11.86 | 8.35 | 10.38 | 17.32 | 1.53 | 1.03 | 1.23 | 1.48 | 15.28 |
| M12 | 9.55 | 4.59 | 1.86 | 1.52 | 12.31 | 3.41 | 99.01 | 0.00 | 23.40 | 19.28 | 2.15 | 8.12 | 22.86 | 1.90 | 1.61 | 2.67 | 3.39 | 30.79 |
| M13 | 74.07 | 91.13 | 6.12 | 10.83 | 12.33 | 16.12 | 45.77 | 15.12 | 9.30 | 7.23 | 0.79 | 2.56 | 9.97 | 6.10 | 3.40 | 2.88 | 1.28 | 18.70 |
| M14 | 5.95 | 3.33 | 0.52 | 14.87 | 25.48 | 38.93 | 67.85 | 60.92 | 23.77 | 65.88 | 87.11 | 14.91 | 56.16 | 5.14 | 1.10 | 3.56 | 2.32 | 21.08 |
| M15 | 66.28 | 87.58 | 3.43 | 22.99 | 35.87 | 52.06 | 79.61 | 77.53 | 38.72 | 42.77 | 46.04 | 27.79 | 54.87 | 13.43 | 3.01 | 5.96 | 2.66 | 24.71 |
| M16 | 8.32 | 16.01 | 7.29 | 3.20 | 39.84 | 13.58 | 0.05 | 14.40 | 30.13 | 24.57 | 1.48 | 14.04 | 13.10 | 12.06 | 9.05 | 12.29 | 8.26 | 51.70 |
| M17 | 8.16 | 4.63 | 0.58 | 10.86 | 5.92 | 43.68 | 97.95 | 12.52 | 26.39 | 19.82 | 1.26 | 8.96 | 51.01 | 9.45 | 1.95 | 2.47 | 1.88 | 3.89 |
| M18 | 6.11 | 4.41 | 0.22 | 7.08 | 6.85 | 24.11 | 81.44 | 4.13 | 14.34 | 17.21 | 0.75 | 4.94 | 30.74 | 2.64 | 1.67 | 4.33 | 1.70 | 100.00 |
| M19 | 9.17 | 6.90 | 0.81 | 59.10 | 81.37 | 14.01 | 41.30 | 27.93 | 14.07 | 20.15 | 7.49 | 9.42 | 16.03 | 7.03 | 1.91 | 9.74 | 4.72 | 29.09 |
| M20 | 8.12 | 13.55 | 4.99 | 6.32 | 3.52 | 18.58 | 36.98 | 26.19 | 47.72 | 23.54 | 2.38 | 4.80 | 15.91 | 79.73 | 18.94 | 9.74 | 2.48 | 16.60 |
| M21 | 5.92 | 4.37 | 0.37 | 2.65 | 2.57 | 5.20 | 65.39 | 35.32 | 38.83 | 99.71 | 36.92 | 3.10 | 26.20 | 12.51 | 2.74 | 6.72 | 1.94 | 25.82 |

M1. Anxiety + Depression
M2. Asthma + Depression + Anxiety
M3. Anxiety + Asthma
M4. SMI + Depression + Anxiety
M5. Asthma + Hypertension + Cancer
M6. Hypertension + Depression + Anxiety
M7. Hypertension + Diabetes + Mental
M8. Mental + Diabetes + Cancer
M9. Asthma-predominant Respiratory + Hypertension
M10. Hypertension + Diabetes
M11. CHD-predominant Cardiovascular + Diabetes
M12. Hypertension + Cancer + Renal
M13. Mental + Physical Long-term Conditions
M14. Cardiovascular + Cardiometabolic + Renal (Complex)
M15. Mental + Cardiovascular + Cardiometabolic + Renal + Respiratory (Complex)
M16. Cancer + COPD + Cardiovascular
M17. Cardiometabolic + Renal
M18. Cancer + Physical Long-term Conditions
M19. COPD-predominant Respiratory + Cardiovascular
M20. Dementia-predominant Neurological + Cardiovascular
M21. AF-predominant Cardiovascular

**b**

| | Anx | Dep | SMI | Ast | COPD | Diab | Hyp | CHD | Stroke | AF | HF | PAD | CKD | Dem | Park | Ost | RA | Can |
|---|---|---|---|---|---|---|---|---|---|---|---|---|---|---|---|---|---|---|
| F1 | 100.00 | 100.00 | 1.94 | 2.19 | 0.83 | 1.26 | 1.52 | 0.47 | 0.40 | 0.18 | 0.02 | 0.08 | 0.39 | 0.05 | 0.05 | 1.02 | 0.51 | 2.66 |
| F2 | 74.41 | 81.43 | 2.10 | 99.96 | 1.90 | 1.89 | 0.02 | 0.24 | 0.34 | 0.13 | 0.03 | 0.05 | 0.29 | 0.01 | 0.02 | 0.29 | 0.40 | 1.21 |
| F3 | 100.00 | 0.00 | 0.00 | 100.00 | 0.00 | 0.00 | 0.00 | 0.00 | 0.00 | 0.02 | 0.00 | 0.00 | 0.00 | 0.00 | 0.00 | 0.00 | 0.00 | 0.00 |
| F4 | 24.71 | 43.91 | 29.10 | 48.60 | 3.96 | 12.88 | 31.18 | 1.27 | 3.48 | 1.14 | 1.38 | 0.54 | 2.67 | 0.06 | 0.06 | 3.59 | 5.60 | 17.43 |
| F5 | 0.00 | 100.00 | 0.00 | 100.00 | 0.00 | 0.00 | 0.00 | 0.00 | 0.00 | 0.00 | 0.00 | 0.00 | 0.00 | 0.00 | 0.00 | 0.00 | 0.00 | 0.00 |
| F6 | 100.00 | 100.00 | 79.98 | 27.26 | 0.92 | 19.10 | 0.00 | 0.00 | 0.00 | 0.00 | 0.00 | 0.00 | 0.00 | 0.00 | 0.00 | 0.00 | 0.00 | 0.00 |
| F7 | 57.53 | 71.65 | 2.97 | 22.57 | 2.76 | 17.81 | 100.00 | 3.93 | 3.50 | 1.03 | 0.14 | 0.65 | 5.49 | 0.16 | 0.12 | 1.19 | 1.77 | 6.81 |
| F8 | 32.26 | 57.52 | 19.68 | 1.16 | 13.51 | 17.87 | 0.08 | 7.16 | 8.91 | 4.42 | 1.08 | 1.85 | 9.65 | 3.46 | 1.79 | 7.41 | 9.59 | 43.36 |
| F9 | 34.85 | 30.92 | 3.43 | 100.00 | 22.87 | 14.99 | 3.35 | 3.45 | 3.22 | 1.75 | 0.34 | 0.71 | 3.87 | 0.23 | 0.24 | 5.94 | 5.84 | 20.29 |
| F10 | 8.25 | 6.17 | 1.58 | 12.14 | 3.67 | 49.38 | 98.58 | 16.48 | 12.60 | 6.26 | 0.29 | 3.84 | 36.09 | 5.49 | 0.70 | 3.18 | 3.30 | 11.52 |
| F11 | 28.73 | 41.83 | 1.92 | 28.05 | 19.77 | 36.35 | 40.11 | 70.92 | 20.28 | 18.09 | 20.54 | 13.63 | 15.05 | 0.82 | 0.26 | 4.91 | 4.17 | 5.77 |
| F12 | 11.22 | 13.03 | 0.42 | 99.22 | 27.04 | 11.14 | 65.31 | 10.46 | 5.05 | 5.56 | 2.24 | 1.41 | 11.95 | 2.72 | 0.51 | 16.52 | 3.73 | 14.48 |
| F13 | 14.32 | 14.20 | 0.68 | 5.70 | 9.52 | 3.30 | 51.10 | 10.88 | 10.84 | 7.49 | 0.95 | 2.51 | 13.93 | 3.53 | 2.24 | 94.87 | 9.83 | 25.81 |
| F14 | 84.51 | 96.31 | 5.55 | 13.13 | 9.63 | 10.55 | 47.90 | 8.36 | 7.63 | 4.72 | 0.49 | 1.26 | 15.33 | 8.02 | 2.01 | 13.83 | 2.26 | 14.78 |
| F15 | 73.80 | 93.13 | 3.67 | 34.27 | 31.27 | 46.01 | 84.54 | 57.33 | 33.00 | 38.05 | 33.42 | 15.70 | 58.75 | 14.30 | 1.81 | 24.24 | 4.27 | 20.83 |
| F16 | 8.02 | 5.44 | 0.42 | 12.58 | 14.47 | 29.53 | 66.53 | 55.92 | 27.13 | 55.59 | 50.22 | 10.83 | 48.70 | 6.71 | 0.78 | 14.08 | 4.03 | 14.66 |
| F17 | 16.58 | 14.76 | 1.05 | 4.46 | 6.48 | 5.06 | 97.28 | 15.06 | 14.42 | 11.44 | 0.01 | 2.95 | 24.65 | 1.88 | 1.34 | 9.78 | 4.47 | 40.79 |
| F18 | 12.63 | 17.24 | 4.69 | 6.38 | 3.95 | 11.38 | 54.31 | 14.94 | 30.75 | 14.11 | 0.88 | 1.99 | 28.36 | 97.62 | 5.17 | 23.94 | 2.84 | 13.20 |

F1. Anxiety + Depression
F2. Asthma + Depression + Anxiety
F3. Anxiety + Asthma
F4. Mental + Asthma + Hypertension
F5. Depression + Asthma
F6. Anxiety + Depression + SMI
F7. Hypertension + Depression + Anxiety
F8. Mental + Cancer + Diabetes
F9. Asthma-predominant Respiratory + Mental
F10. Cardiometabolic + Renal
F11. CHD-predominant Cardiovascular + Diabetes + Mental
F12. Asthma-predominant Respiratory + Hypertension
F13. Osteoporosis-predominant Musculoskeletal + Hypertension + Cancer
F14. Mental + Physical Long-term Conditions
F15. Mental + Cardiovascular + Cardiometabolic + Renal + Respiratory + Musculoskeletal (Complex)
F16. Cardiovascular + Cardiometabolic + Renal (Complex)
F17. Hypertension + Cancer + Renal
F18. Dementia-predominant Neurological + Cardiovascular + Musculoskeletal

**Figure S5: Condition prevalence for each multimorbidity profile. a,** Males. **b,** Females. Condition abbreviations: anxiety (Anx), depression (Dep), serious mental illness (SMI), asthma (Ast), chronic obstructive pulmonary disease (COPD), diabetes (Diab), hypertension (Hyp), coronary heart disease (CHD), stroke or transient ischaemic attack (Stroke), atrial fibrillation (AF), heart failure (HF), peripheral arterial disease (PAD), chronic kidney disease (CKD), osteoporosis (Ost), rheumatoid arthritis (RA), cancer excluding non-melanoma skin cancers (Can).

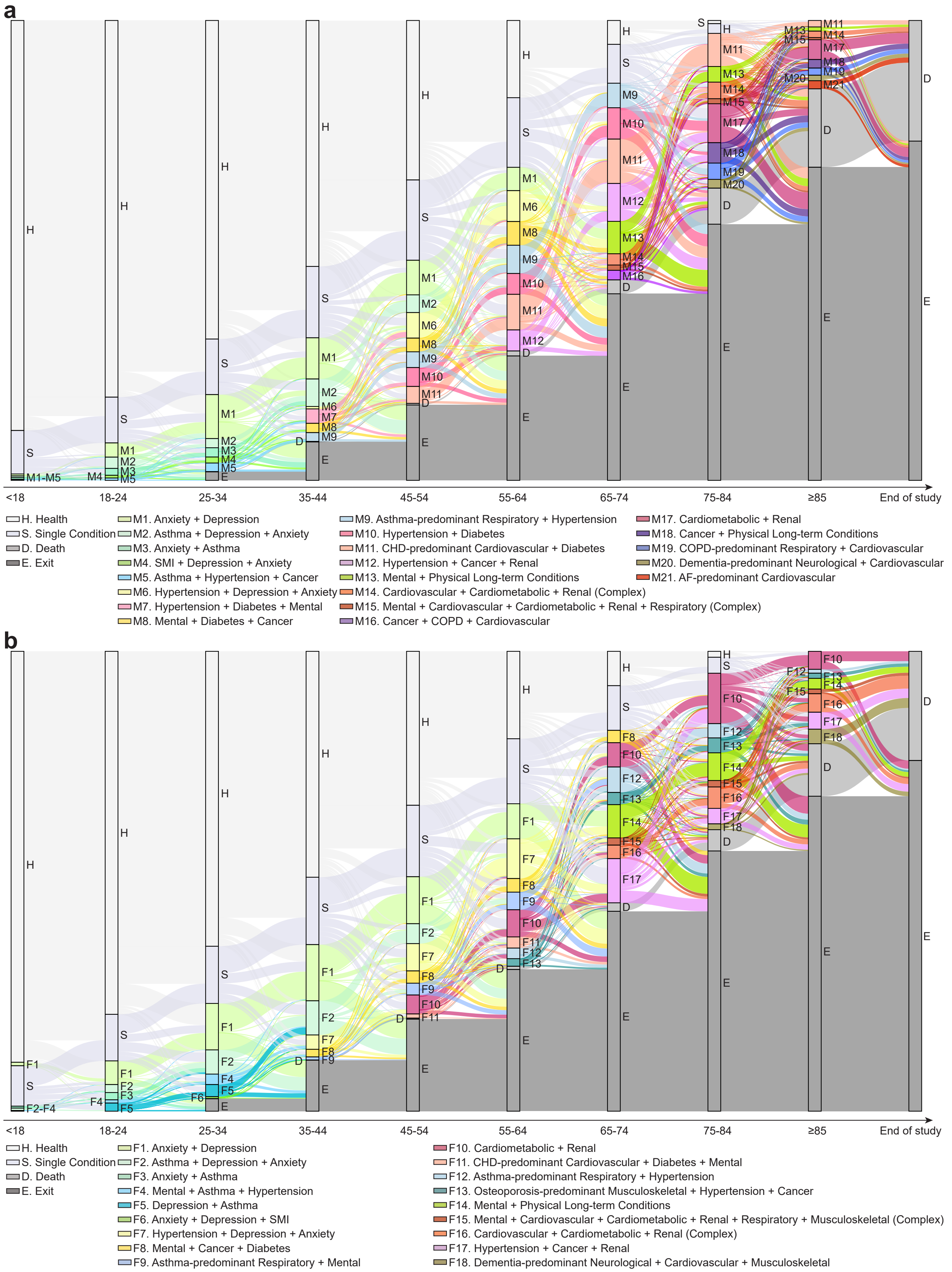

**Figure S6: The Complete multimorbidity trajectories over the life course. a,** Males. **b,** Females. The Sankey diagrams illustrate transitions of the 3.3 million individuals in the primary study cohort between multimorbidity profiles across different age bands, including transitions from individuals with no or single conditions into multimorbidity profiles. The colour coding follows the same scheme as in Fig. 3.

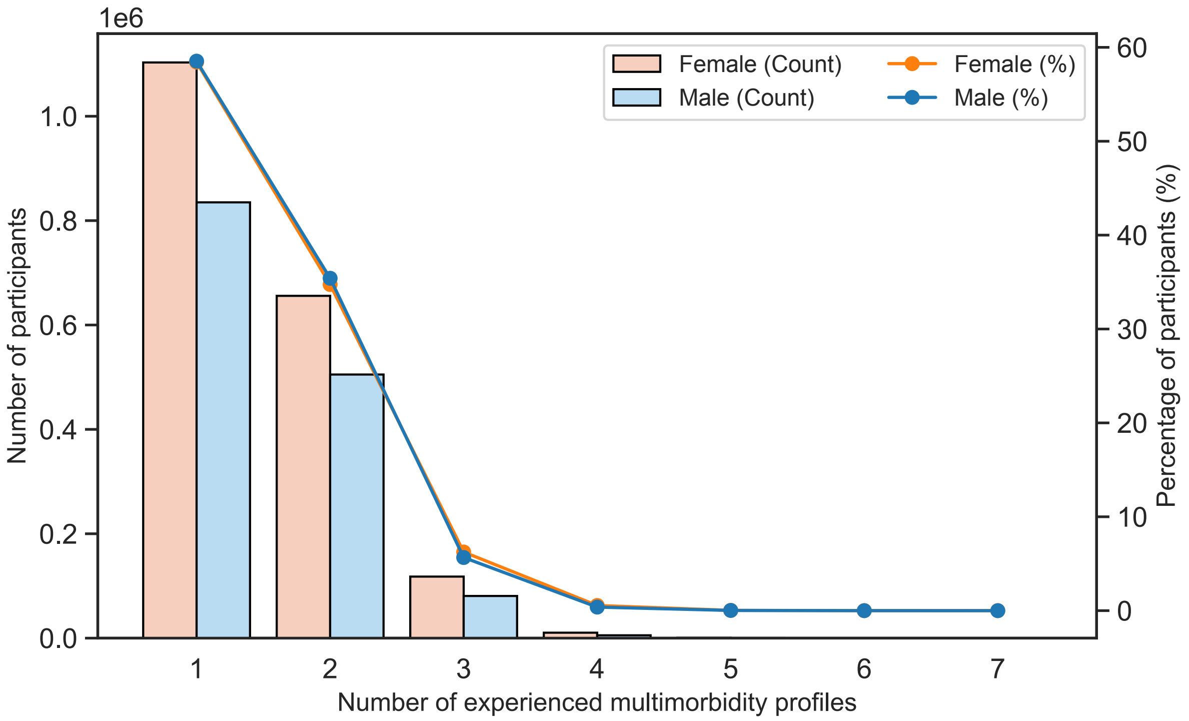

**Figure S7: Distribution of Individuals by Number of Experienced Multimorbidity Profiles.** This figure illustrates the distribution of individuals based on the number of multimorbidity profiles experienced, derived from the multimorbidity trajectories shown in Fig. 3. The bars represent the number of individuals in each group, stratified by sex, whereas the overlaid lines depict the corresponding percentages of the 3.3 million primary study cohort.

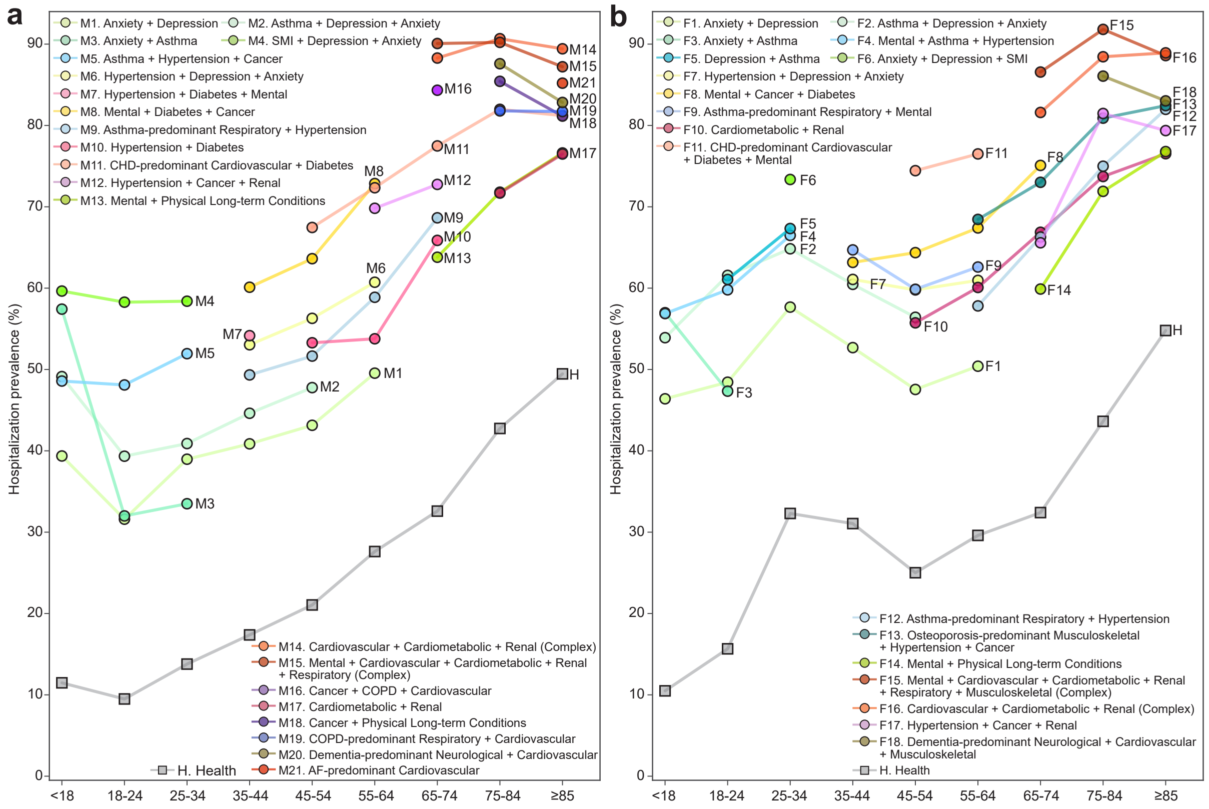

**Figure S8: Multimorbidity burden in terms of hospitalisation prevalence over the life course. a, b**, Hospitalisation prevalence across multimorbidity profiles and age bands in males and females, respectively. Each line represents a multimorbidity profile, with values calculated within profile-specific cohorts and age bands. Hospitalisation prevalence was defined as the proportion of individuals experiencing at least one hospitalisation within the corresponding profile and age band. The healthy cohort is shown in grey for comparison. The colour coding follows the same scheme as in Fig. 3.

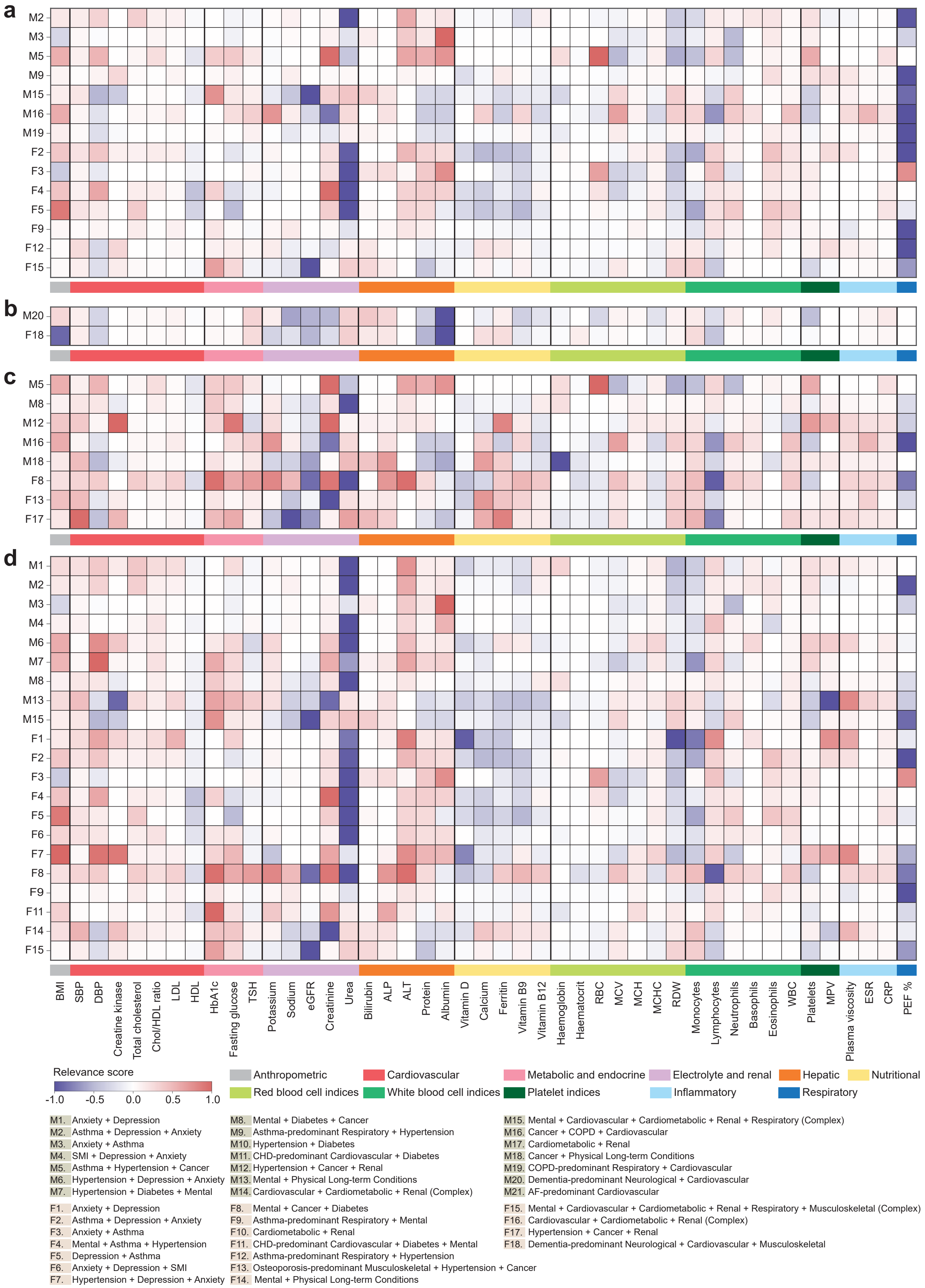

**Figure S9: Relevance of clinical markers across specific groups of multimorbidity profiles (Part I). a,** Respiratory-related profiles, **b** Neurological-related profiles, **c,** Cancer-related profiles, **d,** Mental health-related profiles. The heatmaps show the clinical relevance of each marker (columns) to each multimorbidity profile (rows), using identical methodology to Fig. 6. Profiles appear in multiple panels when spanning disease systems.

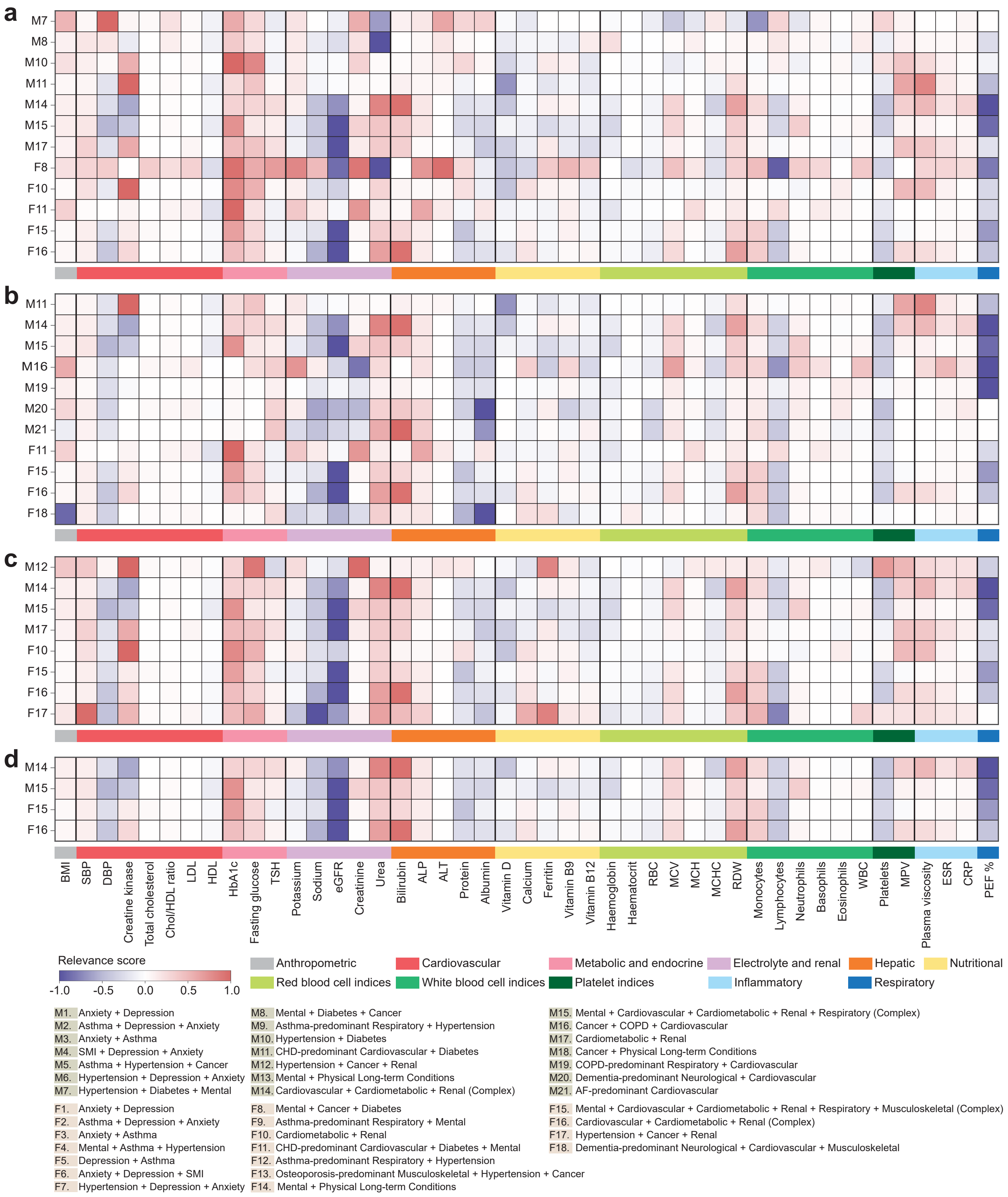

**Figure S10: Relevance of clinical markers across specific groups of multimorbidity profiles (Part II). a,** Cardiometabolic-related profiles, **b,** Cardiovascular-related profiles, **c** Renal-related profiles, **d,** Complex profiles. The heatmaps show the clinical relevance of each marker (columns) to each multimorbidity profile (rows), using identical methodology to Fig. 6. Profiles appear in multiple panels when spanning disease systems.

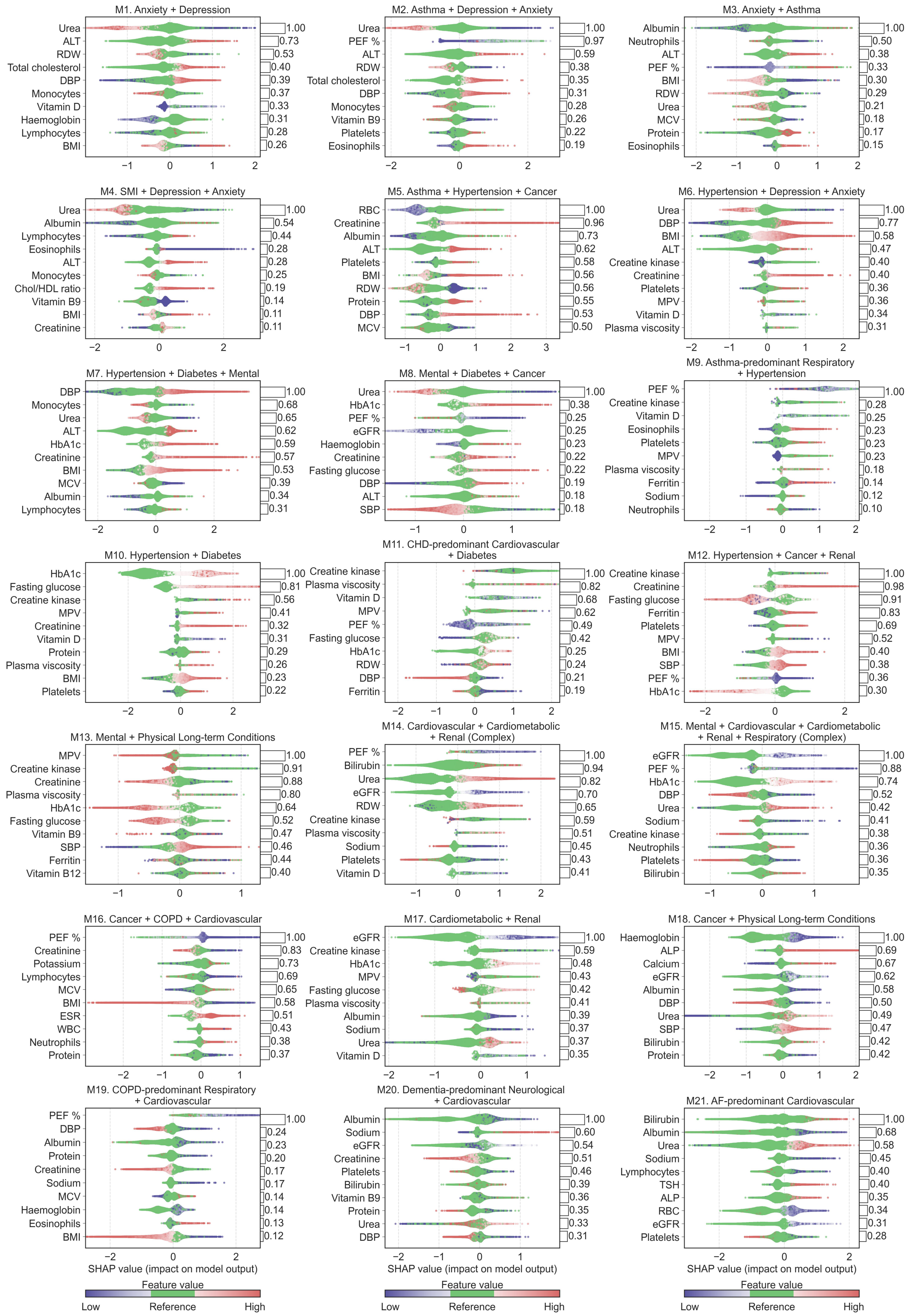

**Figure S11: Beeswarm plots for each multimorbidity profile in males.** In the beeswarm plot, each dot represents an individual, with its horizontal position (SHAP value) indicating the impact of a given marker on the model's prediction. Dots are coloured according to marker values relative to a reference range, with green indicating normal values. When multiple dots share the same $x$ position, they accumulate vertically to reflect density. The bar plots on the right display the relevance scores of corresponding markers based on the positive SHAP contributions of abnormally high or low marker values, which correspond to the relevance scores in Fig. 6a. Top ten important markers for each profile are shown.

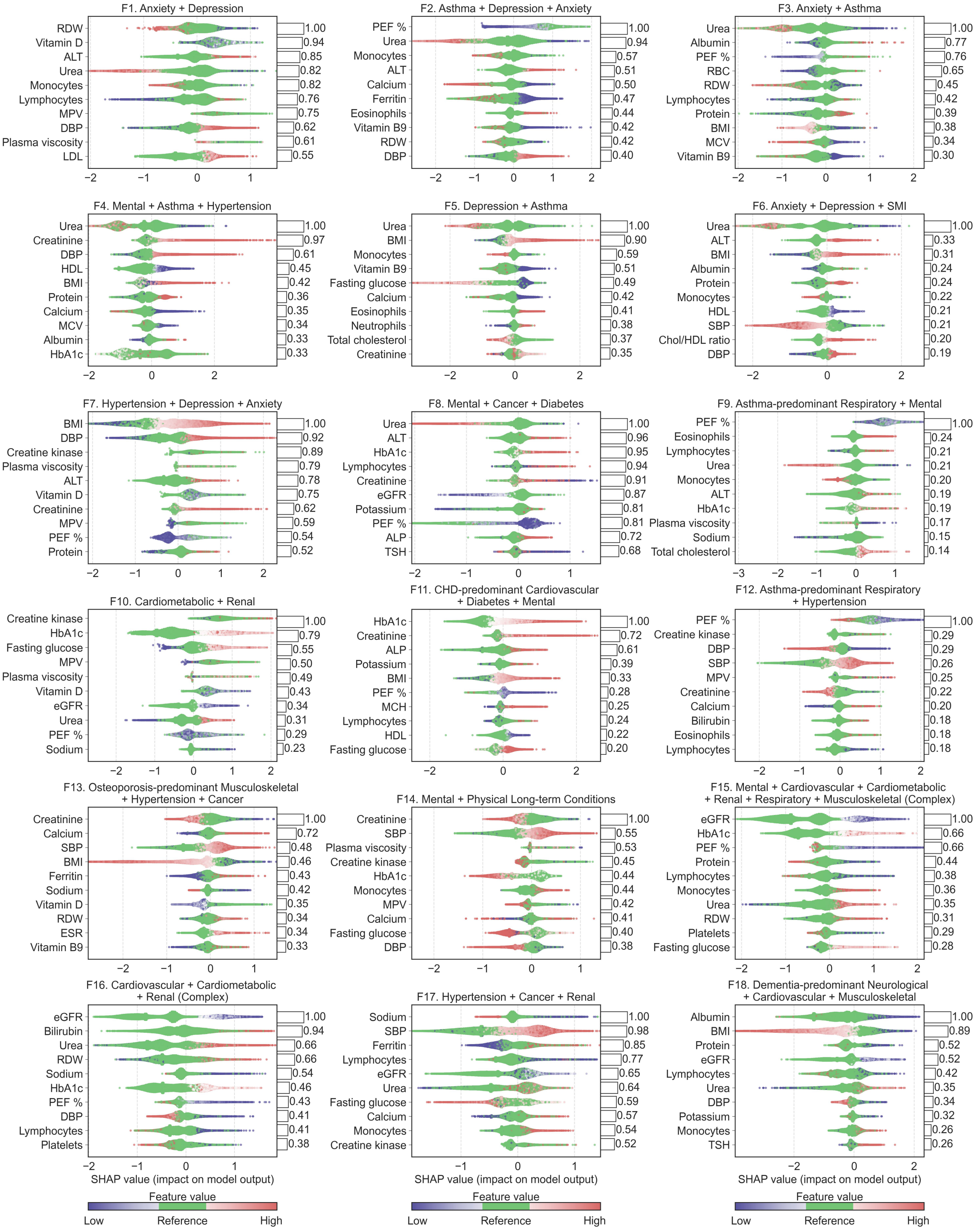

**Figure S12: Beeswarm plots for each multimorbidity profile in females.** In the beeswarm plot, each dot represents an individual, with its horizontal position (SHAP value) indicating the impact of a given marker on the model's prediction. Dots are coloured according to marker values relative to a reference range, with green indicating normal values. When multiple dots share the same $x$ position, they accumulate vertically to reflect density. The bar plots on the right display the relevance scores of corresponding markers based on the positive SHAP contributions of abnormally high or low marker values, which correspond to the relevance scores in Fig. 6b. Top ten important markers for each profile are shown.

### Supplementary Tables

**Table S1: Characteristics of the study population by socioeconomic deprivation, ethnicity, and geographic regions.**

| | Socioeconomic deprivation, $n$ (%) | | | | | |
|---|---|---|---|---|---|---|
| **Sex** | IMD 1 (least deprived) | IMD 2 | IMD 3 | IMD 4 | IMD 5 (most deprived) | Unknown |
| Male | 264,798 (18.56) | 277,660 (19.46) | 274,229 (19.22) | 298,320 (20.91) | 310,289 (21.75) | 1,407 (0.10) |
| Female | 353,772 (18.74) | 368,195 (19.50) | 364,930 (19.33) | 393,674 (20.85) | 405,635 (21.49) | 1,743 (0.09) |
| | **Ethnicity,** $n$ (%) | | | | | |
| **Sex** | White | Black | South Asian | Mixed | Other | Unknown |
| Male | 1,048,953 (73.52) | 39,141 (2.74) | 53,293 (3.74) | 12,008 (0.84) | 24,563 (1.72) | 248,745 (17.44) |
| Female | 1,409,930 (74.68) | 50,483 (2.67) | 56,264 (2.98) | 17,913 (0.95) | 27,168 (1.44) | 326,191 (17.28) |
| | **Geographic regions,** $n$ (%) | | | | | |
| **Sex** | London | East of England | South East | South West | West Midlands | East Midlands |
| Male | 217,744 (15.26) | 58,483 (4.10) | 243,261 (17.05) | 203,710 (14.28) | 244,212 (17.12) | 33,069 (2.32) |
| Female | 276,972 (14.67) | 78,770 (4.17) | 333,153 (17.65) | 270,105 (14.31) | 320,847 (16.99) | 44,590 (2.36) |
| **Sex** | Yorkshire and the Humber | North West | North East | Unknown | | |
| Male | 62,332 (4.37) | 281,880 (19.76) | 62,305 (4.37) | 19,707 (1.38) | | |
| Female | 81,878 (4.34) | 372,362 (19.72) | 82,780 (4.39) | 26,492 (1.40) | | |

IMD, index of multiple deprivation.

**Table S2: Units and reference ranges of clinical markers used in the study.**

| Clinical marker | Unit | Reference range[a] |
|---|---|---|
| ALP (Alkaline phosphatase) | U/L | 30-130 |
| ALT (Alanine aminotransferase) | U/L | 0-50/0-35 |
| Albumin | g/L | 35-50 |
| BMI (Body mass index) | $kg/m^2$ | 18.5-24.9 |
| Basophils | $\times 10^9$/L | 0-0.1 |
| Bilirubin | umol/L | 0-21 |
| CRP (C-reactive protein) | mg/L | 0-5 |
| Calcium | mmol/L | 2.2-2.6 |
| Chol/HDL ratio (Total cholesterol:HDL cholesterol ratio) | ratio | 0.0-4.0 |
| Creatinine | umol/L | 59-104/45-84 |
| Creatine kinase | U/L | 40-320/25-200 |
| DBP (Diastolic blood pressure) | mmHg | 60-80 |
| ESR (Erythrocyte sedimentation rate) | mm/hr | 0-14/0-20 |
| Eosinophils | $\times 10^9$/L | 0.1-0.4/0.0-0.5 |
| eGFR (Estimated glomerular filtration rate) | $mL/min/1.73m^2$ | $\geq$60 |
| Fasting glucose | mmol/L | 3.9-5.4 |
| Ferritin | ug/L | 30-340/30-310 |
| HDL (High-density lipoprotein cholesterol) | mmol/L | $\geq$ 1.0/ $\geq$ 1.2 |
| Haematocrit | ratio | 0.40-0.54/0.37-0.47 |
| Haemoglobin | g/L | 130-180/115-165 |
| HbA1c | ug/L | 0-42 |
| LDL (Low-density lipoprotein cholesterol) | mmol/L | 0.0-3.0 |
| Lymphocytes | $\times 10^9$/L | 1.0-4.0 |
| MCH (Mean corpuscular haemoglobin) | pg | 27-32 |
| MCHC (Mean corpuscular haemoglobin concentration) | g/L | 315-345 |
| MCV (Mean corpuscular volume) | fL | 83-101 |
| MPV (Mean platelet volume) | fL | 7.2-11.7 |
| Monocytes | $\times 10^9$/L | 0.2-0.8 |
| Neutrophils | $\times 10^9$/L | 1.8-7.5 |
| PEF % (Peak expiratory flow percentage) | % | 80-120 |
| Platelets | $\times 10^9$/L | 140-400 |
| Plasma viscosity | mPA·s | 1.50-1.72 |
| Potassium | mmol/L | 3.5-5.3 |
| Protein | g/L | 60-80 |
| RBC (Red blood cell count) | $\times 10^{12}$/L | 4.5-6.5/3.5-5.8 |
| RDW (Red cell distribution width) | % | 11.8-14.5/12.2-16.1 |
| SBP (Systolic blood pressure) | mmHg | 90-120 |
| Sodium | mmol/L | 133-146 |
| TSH (Thyroid stimulating hormone) | mU/L | 0.4-4.0 |
| Total Chol. (Total cholesterol) | mmol/L | 0.0-5.0 |
| Urea | mmol/L | 2.5-7.8 |
| Vitamin B9 | ng/mL | 3-20 |
| Vitamin B12 | pg/mL | 180-1000 |
| Vitamin D | nmol/L | $\geq$50 |
| WBC (White blood cell count) | $\times 10^9$/L | 3.6-11.0 |

[a] Reference ranges for clinical markers are provided as male/female if there is a sex difference; otherwise, a single range is reported.

**Table S3: Population distribution, prevalence, mean number of conditions among multimorbidity profiles in males.**

| Profile | <18 | 18–24 | 25–34 | 35–44 | 45–54 | 55–64 | 65–74 | 75–84 | ≥85 |
|---|---|---|---|---|---|---|---|---|---|
| M1 | 5,581 (0.39)<br>2.00 (0.07) | 44,091 (3.09)<br>2.01 (0.10) | 136,314 (9.74)<br>2.28 (0.47) | 127,769 (9.78)<br>2.09 (0.29) | 107,462 (9.05)<br>2.17 (0.37) | 72,265 (7.05)<br>2.43 (0.59) | | | |
| M2 | 4,694 (0.33)<br>2.32 (0.47) | 34,927 (2.45)<br>2.49 (0.50) | 28,506 (2.04)<br>2.01 (0.08) | 85,303 (6.53)<br>2.55 (0.65) | 54,913 (4.62)<br>2.63 (0.71) | | | | |
| M3 | 5,904 (0.41)<br>2.00 (0.00) | 21,471 (1.50)<br>2.00 (0.00) | 28,737 (2.05)<br>2.01 (0.15) | | | | | | |
| M4 | 835 (0.06)<br>2.26 (0.51) | 8,314 (0.58)<br>2.40 (0.60) | 18,723 (1.34)<br>2.50 (0.66) | | | | | | |
| M5 | 2,744 (0.19)<br>2.05 (0.24) | 7,364 (0.52)<br>2.16 (0.45) | 26,717 (1.91)<br>2.23 (0.55) | | | | | | |
| M6 | | | | 7,591 (0.58)<br>3.06 (0.24) | 79,206 (6.67)<br>3.01 (0.97) | 95,803 (9.34)<br>3.16 (1.06) | | | |
| M7 | | | | 44,577 (3.41)<br>2.44 (0.84) | | | | | |
| M8 | | | | 28,949 (2.22)<br>2.20 (0.58) | 42,408 (3.57)<br>2.42 (0.89) | 73,575 (7.18)<br>2.91 (1.35) | | | |
| M9 | | | | 27,402 (2.10)<br>2.19 (0.46) | 48,936 (4.12)<br>2.32 (0.64) | 87,750 (8.56)<br>2.57 (0.89) | 76,176 (9.47)<br>2.93 (1.05) | | |
| M10 | | | | | 58,582 (4.93)<br>2.18 (0.49) | 64,479 (6.29)<br>2.31 (0.50) | 97,239 (12.08)<br>2.82 (0.91) | | |
| M11 | | | | | 52,434 (4.41)<br>2.68 (1.00) | 110,503 (10.78)<br>2.97 (1.15) | 137,599 (17.10)<br>3.08 (1.06) | 102,693 (19.74)<br>3.38 (1.13) | 20,194 (9.52)<br>3.26 (1.21) |
| M12 | | | | | | 65,339 (6.37)<br>2.45 (0.83) | 117,636 (14.62)<br>2.50 (0.78) | | |
| M13 | | | | | | | 100,473 (12.48)<br>3.13 (1.18) | 48,466 (9.32)<br>3.59 (1.37) | 12,882 (6.07)<br>3.99 (1.42) |
| M14 | | | | | | | 35,033 (4.35)<br>4.57 (1.51) | 51,548 (9.91)<br>4.94 (1.57) | 20,132 (9.49)<br>5.86 (1.34) |
| M15 | | | | | | | 16,411 (2.04)<br>6.61 (1.23) | 15,543 (2.99)<br>7.04 (1.30) | 6,610 (3.12)<br>7.03 (1.34) |
| M16 | | | | | | | 29,650 (3.68)<br>2.79 (1.01) | | |
| M17 | | | | | | | | 120,804 (23.23)<br>2.90 (1.07) | 61,335 (28.92)<br>3.53 (1.24) |
| M18 | | | | | | | | 63,259 (12.16)<br>3.14 (1.04) | 26,449 (12.47)<br>3.10 (0.91) |
| M19 | | | | | | | | 50,633 (9.73)<br>3.46 (1.42) | 22,208 (10.47)<br>3.93 (1.52) |
| M20 | | | | | | | | 27,083 (5.21)<br>3.48 (1.40) | 17,109 (8.07)<br>3.28 (1.26) |
| M21 | | | | | | | | | 25,150 (11.86)<br>3.76 (1.28) |

Each cell presents the population (prevalence, %), mean number of conditions per individual (SD) for the multimorbidity profile within the corresponding age band.

#### Table S4: Population distribution, prevalence, mean number of conditions among multimorbidity profiles in females.

| Profile | <18 | 18–24 | 25–34 | 35–44 | 45–54 | 55–64 | 65–74 | 75–84 | ≥85 |
|---|---|---|---|---|---|---|---|---|---|
| F1 | 13,898 (0.74)<br>2.01 (0.11) | 96,994 (5.14)<br>2.02 (0.16) | 190,841 (10.39)<br>2.03 (0.16) | 230,597 (13.75)<br>2.12 (0.33) | 192,927 (12.81)<br>2.15 (0.35) | 144,093 (11.14)<br>2.38 (0.53) | | | |
| F2 | 10,154 (0.54)<br>2.33 (0.47) | 32,940 (1.74)<br>3.00 (0.05) | 99,301 (5.41)<br>2.61 (0.49) | 140,380 (8.37)<br>2.55 (0.61) | 81,829 (5.43)<br>2.76 (0.67) | | | | |
| F3 | 7,643 (0.40)<br>2.00 (0.00) | 29,764 (1.58)<br>2.00 (0.02) | 28,737 (2.05)<br>2.01 (0.15) | | | | | | |
| F4 | 2,866 (0.15)<br>2.10 (0.34) | 13,304 (0.70)<br>2.30 (0.62) | 42,915 (2.34)<br>2.33 (0.66) | | | | | | |
| F5 | | 33,473 (1.77)<br>2.00 (0.00) | 49,762 (2.71)<br>2.00 (0.00) | | | | | | |
| F6 | | | 7,717 (0.42)<br>3.27 (0.45) | | | | | | |
| F7 | | | | 58,681 (3.50)<br>2.56 (0.88) | 111,893 (7.43)<br>2.99 (0.91) | 162,979 (12.60)<br>3.17 (1.02) | | | |
| F8 | | | | 31,395 (1.87)<br>2.17 (0.55) | 50,357 (3.34)<br>2.25 (0.63) | 55,919 (4.32)<br>2.39 (0.79) | 50,684 (4.91)<br>2.73 (1.07) | | |
| F9 | | | | 12,377 (0.74)<br>2.13 (0.45) | 48,465 (3.22)<br>2.41 (0.83) | 72,127 (5.58)<br>2.74 (1.03) | | | |
| F10 | | | | | 77,874 (5.17)<br>2.26 (0.56) | 111,683 (8.64)<br>2.32 (0.63) | 99,840 (9.67)<br>2.98 (0.94) | 206,791 (28.24)<br>2.95 (1.05) | 74,026 (19.52)<br>3.41 (1.17) |
| F11 | | | | | 17,024 (1.13)<br>3.46 (1.51) | 45,653 (3.53)<br>3.81 (1.66) | | | |
| F12 | | | | | | 43,423 (3.36)<br>2.49 (0.65) | 104,410 (10.11)<br>2.86 (1.00) | 58,643 (8.01)<br>3.29 (1.28) | 15,773 (4.16)<br>4.54 (1.69) |
| F13 | | | | | | 31,417 (2.43)<br>2.61 (0.91) | 50,240 (4.87)<br>2.72 (0.93) | 61,264 (8.37)<br>2.99 (1.02) | 21,188 (5.59)<br>2.85 (1.06) |
| F14 | | | | | | | 136,349 (13.21)<br>3.16 (0.98) | 113,759 (15.54)<br>3.61 (1.33) | 43,948 (11.59)<br>4.03 (1.40) |
| F15 | | | | | | | 29,669 (2.87)<br>6.14 (1.19) | 26,363 (3.60)<br>7.05 (1.25) | 19,132 (5.04)<br>7.02 (1.33) |
| F16 | | | | | | | 55,508 (5.38)<br>3.88 (1.46) | 87,739 (11.98)<br>4.26 (1.57) | 75,926 (20.02)<br>4.53 (1.48) |
| F17 | | | | | | | 181,436 (17.57)<br>2.47 (0.79) | 63,586 (8.69)<br>3.06 (0.95) | 69,518 (18.33)<br>3.08 (1.12) |
| F18 | | | | | | | | 23,912 (3.27)<br>3.44 (1.35) | 59,797 (15.76)<br>3.45 (1.19) |

Each cell presents the population (prevalence, %), mean number of conditions per individual (SD) for the multimorbidity profile within the corresponding age band.
